# Aspirin improves both reactivity and durability of type-I interferon signaling to achieve functional cure of chronic hepatitis B

**DOI:** 10.1101/2024.06.14.24308555

**Authors:** Ying Miao, Yukang Yuan, Yuanmei Chen, Jin Liu, Fan Huang, Tingting Zhang, Renxia Zhang, Qian Zhao, Qun Cui, Wanying Tian, Wei He, Yibo Zuo, Zhijin Zheng, Zhenglan Zhao, Ming Li, Feng Qian, Li Zhu, Chuanwu Zhu, Hui Zheng

## Abstract

Type-I interferon (IFN-I) is currently the only drug for achieving a functional cure of chronic hepatitis B-virus (HBV) infection that is defined as HBsAg loss. However, the IFN-I-mediated functional cure rate is extremely low thus far. Previous studies demonstrated that IFN-I-induced degradation of IFN-I receptor-1 (IFNAR1) restricts the reactivity of IFN-I signaling. Here, we further reveal that IRF9 de-phosphorylation inhibits the durability of IFN-I signaling. We found that IRF9-Tyr112 phosphorylation is critical for IRF9 binding to the promoters of interferon-stimulated genes (ISGs), while PTP1B induces IRF9 de-phosphorylation and therefore attenuates IFN-I signaling durability and ISGs expression. Interestingly, we found that Aspirin can both rescue IRF9 phosphorylation and inhibit IFNAR1 degradation, thus remolding IFN-I signaling. Importantly, the functional cure rate after the IFN-I and Aspirin combination (IA) therapy reached over 86% (13/15). This study reveals the IA therapy as an effective therapeutic way for achieving a chronic HBV functional cure.

## Introduction

The hepatitis B virus (HBV) was first discovered in 1963 due to the identification of its surface antigen (HBsAg) (Millman et al., 1970). HBV infection has been a major global health burden with around 296 million people infected in 2019 and 1.5 million new infections each year worldwide (Jeng et al., 2023). HBV infection could lead to acute or chronic hepatitis, which may develop liver cirrhosis and hepatocellular carcinoma (HCC). It has been reported that hepatitis viruses account for over 70% of all HCC cases worldwide (Forner et al., 2018). Current FDA-approved therapies for chronic HBV infection include type-I interferons (IFN-I) and nucleos(t)ide analogues (NA). NA can lead to high rates of virologic suppression, but requires a lifelong duration of maintenance therapy and cannot achieve a functional cure (the therapeutic end point) of chronic HBV infection, which is defined as HBsAg loss in serum (van Bömmel et al., 2023). Compared to NA, IFN-I is currently the only drug that can achieve a functional cure of chronic HBV infection. However, IFN-I-mediated functional cure rate of chronic HBV infection is still around 10% thus far (Ignat et al., 2024; Lam and Lim, 2024; Tang et al., 2022). Recently, the World Health Organization (WHO) has raised a goal for HBV elimination by 2030. Thus, it is of paramount significance to improve the IFN-I-mediated functional cure rate of HBV infection.

IFN-I can suppress HBV DNA replication, which in turn inhibits liver inflammation and lowers the risk of liver cirrhosis and HCC. In addition, IFN-I can regulate cytotoxic T-cell immune responses in HBV patients, which are associated with IFN-I-induced HBsAg loss, the key marker of an HBV functional cure (Rehermann et al., 1996). Importantly, IFN-I promotes the degradation of covalently closed circular DNA (cccDNA) of HBV and decreases cccDNA transcription (Lucifora et al., 2014). Despite these multiple effects, IFN-I-mediated HBV functional cure is extremely low until now. It has been reported that HBsAg loss is around 7% of both hepatitis-B e Ag (HBeAg) positive and negative patients with a year of IFN-I treatment (Flink et al., 2006). In addition, HBsAg loss increases from 2%–3% at the end of 48-week IFN-I treatment to 8%–14% after 5 years post-treatment follow-up (Buster et al., 2008; Marcellin et al., 2013). This treatment status is far lower than people’s expectations, suggesting that IFN-I antiviral activity could be largely compromised in the treatment of HBV infection.

IFN-I (IFNα/β) exerts its antiviral activity through the Janus kinase (JAK)-signal transducer and activator of transcription (STAT) signaling pathway. IFN-I first binds with the IFN-I receptors (IFNAR1 and IFNAR2) that are ubiquitously expressed on cytoplasmic membranes of almost all types of cells. Upon IFN-I binding, JAK family members (JAK1 and Tyk2) are activated and induce STAT1 phosphorylation. STAT1 and STAT2, together with interferon regulatory factor 9 (IRF9), form the interferon-stimulated gene factor 3 (ISGF3) complex (Stark and Darnell, 2012). IRF9 as the DNA binding subunit of ISGF3 can directly bind with the IFN-stimulated response element (ISRE) located in the promoters of interferon-stimulated genes (ISGs) (Nan et al., 2018). Subsequently, nuclear ISGF3 complex induces transcriptional expression of ISGs, which execute IFN-I antiviral activity (Zuo et al., 2020; Zuo et al., 2022).

Previous studies from the Fuchs team and ours have intensively demonstrated that IFN-I-induced ubiquitination and degradation of IFN-I receptor IFNAR1 attenuates IFN-I signaling (Guo et al., 2019; Kumar et al., 2004; Kumar et al., 2003; Zheng et al., 2011b). IFNAR1 degradation is considered to be an important negative feedback regulation of IFN-I signaling, thus attenuating the reactivity of IFN-I signaling and the efficacy of IFN-I antiviral therapy. Our further studies suggest that besides IFN-I-induced IFNAR1 degradation, it is very worth exploring the other mechanisms that severely restrict IFN-I signaling and therapeutic efficacy. In this study, we revealed IFN-I-induced IRF9 de-phosphorylation as another crucial negative regulation of IFN-I signaling, which disrupts the durability of IFN-I signaling. Interestingly, we found that Aspirin can inhibit IFNAR1 protein degradation, while the metabolite of Aspirin in the body, Salicylate, can inhibit IRF9 de-phosphorylation. Importantly, Aspirin administration *in vivo* dramatically improves a functional cure of chronic HBV infection by enhancing both the reactivity and durability of IFN-I signaling.

## Results

### Protein tyrosine de-phosphorylation attenuates the durability of IFN-I signaling

Previous studies revealed that IFN-I stimulates IFNAR1 phosphorylation at Ser535, which in turn recruits a ubiquitin E3 ligase β-TrCP to induce IFNAR1 ubiquitination and degradation (Kumar et al., 2004; Zheng et al., 2011a; Zheng et al., 2011b). Further studies demonstrated that β-TrCP-mediated IFNAR1 degradation is critically regulated by PARP11 and leads to IFN-I antiviral efficiency loss (Guo et al., 2019) (Fig. 1A). Consistently, IFN-I-induced expression of antiviral ISGs showed a gradual decay in cells with long-time IFN-I treatment (Fig. S1A). Similarly, we observed IFNAR1 downregulation in the peripheral blood mononuclear cells (PBMCs) of chronic HBV infection patients with Pegylated-IFN (Peg-IFN) treatment (Fig. 1B). Importantly, ISG expression in these patients at 6 h after Peg-IFN treatment showed only slight and even no upregulation (Fig. 1C). Based on these findings, we speculated that maintaining IFNAR1 stability could be able to inhibit IFN-I signaling decay in cells. Given that substituting Ala535 for Ser535 on IFNAR1 (S535A) maintains IFNAR1 stability and enhances IFN-I antiviral activity (Kumar et al., 2003; Zheng et al., 2013; Zheng et al., 2011b), we employed *Ifnar1^-/-^* cells stably expressing wild-type (WT) or S535A of IFNAR1 to analyze IFN-induced ISG transcription during the time course of IFN treatment. This result showed that maintaining IFNAR1 stability still cannot abolish IFN-I signaling decay in IFNAR1-S535A cells, but did significantly enhance the strength of IFN-I signaling (Fig. 1D). These data suggested that certain other mechanisms could mediate the decay of IFN-I signaling in cells.

**Figure 1.**
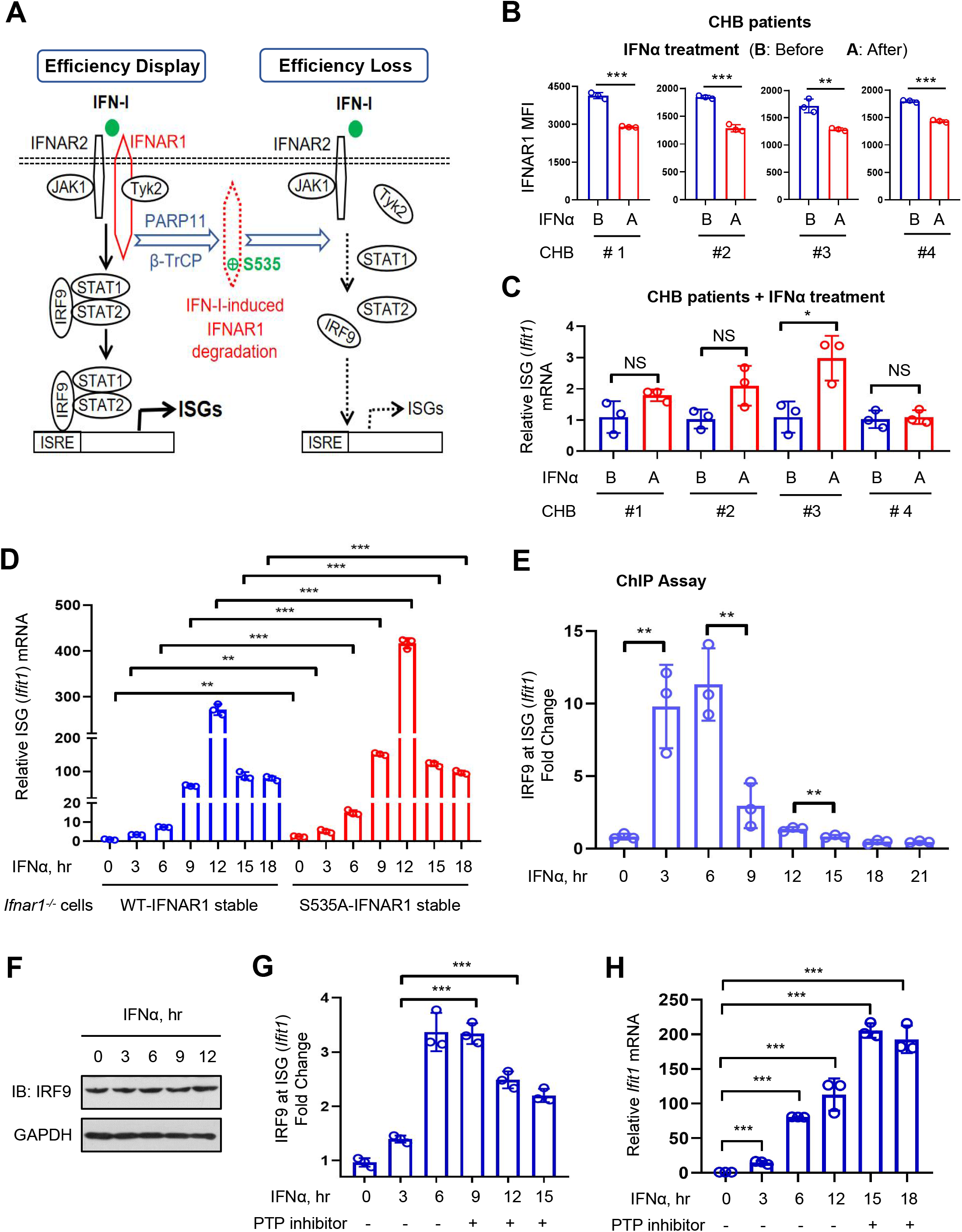
Protein tyrosine de-phosphorylation attenuates the durability of IFN-I signaling. (A) A diagram showing that IFN-I-induced IFNAR1 degradation mediated by PARP11 and β-TrCP leads to a shift in IFN-I antiviral efficiency from “Efficiency Display” to “Efficiency Loss”. (B and C) Flow cytometry analysis of cytoplasmic membrane IFNAR1 levels (B) or RT-qPCR analysis of *Ifit1* mRNA (C) in PBMCs from chronic hepatitis-B (CHB) patients before and 6 h after Pegylated (Peg)-IFN treatment (180 μg each person). B: before treatment; A: after treatment. (D) IFNAR1 (wild-type,WT; S535A mutants) constructs were stably transfected in *Ifnar1^-/-^* HEK293T cells. RT-qPCR was used to analyze *Ifit1* mRNA in these cells treated with IFNα (1,000 IU/ml) for indicated times. (E) ChIP-qPCR analysis of the binding of cellular IRF9 proteins to the ISRE promoters of the representative ISG (*Ifit1*) gene in HEK293T cells treated with IFNα (1,000 IU/ml) for indicated times. (F) Western blot analysis of IRF9 in HT1080 cells treated with IFNα (1,000 IU/ml) for indicated times. (G) ChIP-qPCR analysis of the binding of IRF9 proteins to the ISRE of *Ifit1* in HEK293T cells treated with PTP inhibitor I (10 μM) and (or) IFNα (1,000 IU/ml) as indicated. (H) RT-qPCR analysis of *Ifit1* mRNA in HEK293T cells treated with PTP inhibitor I (10 μM) and (or) IFNα (1,000 IU/ml) as indicated. NS, not significant (p > 0.05), *p < 0.05, **p < 0.01, ***p < 0.001 (two-tailed unpaired Student’s *t*-test). Data are shown as means ± SD of three biological replicates (B-E, G, H), or are representative of three independent experiments (F).

We next sought to delay IFN-I signaling decay to obtain durable ISG transcriptional expression. Given that IRF9-ISRE binding is the base of ISG transcription, we focused on IRF9-mediated binding of the ISGF3 transcription complex to the ISRE of ISG promoters. We found that IFN-I first stimulated IRF9-ISRE binding, which then was gradually reduced with the extension of IFN-I treatment time (Fig. 1E). This showed a similar tendency as IFN-induced ISG expression, despite that there was a difference in the time reaching the peak level, which may be due to continued accumulation of ISG mRNA resulting from the ISG mRNA stability in cells. We further demonstrated that IFN-I treatment did not reduce total IRF9 protein levels (Fig. 1F). Thus, we hypothesized that protein post-translational modifications (PTMs) of IRF9 could affect the IRF9-ISRE binding.

As well known, IFN-I-induced tyrosine phosphorylation is widely present on almost all IFN-I signaling node proteins and plays key roles in activating IFN-I signaling. However, whether IRF9 undergoes tyrosine phosphorylation and how tyro-phosphorylated IRF9 regulates IFN-I signaling remain unknown thus far. Interestingly, we observed that IRF9 has tyrosine phosphorylation modifications in cells, which showed a dramatic reduction during long-time IFN-I treatment in a dose-dependent manner (Fig. S1B and S1C). Based on these results, we speculated that IRF9 tyrosine de-phosphorylation could contribute to the reduction of IRF9-ISRE binding and subsequent ISG transcriptional expression, which lowers the durability of IFN-I signaling. Thus, we utilized a pan-phosphatase inhibitor to inhibit IRF9 de-phosphorylation during IFN-I treatment. We found that the phosphatase inhibitor indeed blocked the downregulation of IRF9 tyrosine phosphorylation (Fig. S1D), and delayed the reduction of IRF9-ISRE binding induced by durable IFN-I treatment (Fig. 1G). Importantly, inhibition of tyrosine phosphorylation significantly delayed the decay of ISG transcriptional expression (Fig. 1H). Taken together, these findings suggested that IRF9 tyrosine phosphorylation could be critical to the durability of IFN-I signaling.

### IRF9 Tyr112 phosphorylation is critical for IRF9-ISRE binding but is reduced with durable IFN-I treatment

To further study IRF9 tyrosine phosphorylation, we first utilized mass spectrometry to identify the tyrosine phosphorylation residue of IRF9. Our results showed only one tyro-phosphorylated residue, Tyr112, on IRF9 proteins (Fig. 2A). Given that IRF9 has eight tyrosine residues in total, we mutated each tyrosine residue to observe IRF9 tyrosine phosphorylation. The results showed that Tyr112 mutation on Flag-GFP-tagged IRF9 (F-G-IRF9) almost abolished its tyrosine phosphorylation (Fig. 2B). Importantly, IRF9-Tyr112 mutation dramatically restricted the binding of IRF9 to the ISRE of ISG promoters (Fig. 2C), and inhibited IFN-I-stimulated IRF9-ISRE binding (Fig. 2C). Consistently, IRF9-Tyr112 mutation significantly decreased not only basal levels of ISG transcription but also IFN-I-induced ISG transcription (Fig. 2D). These findings demonstrated that IRF9-Tyr112 phosphorylation is critical for IRF9 binding to the ISG promoters and subsequent ISG transcription expression.

**Figure 2.**
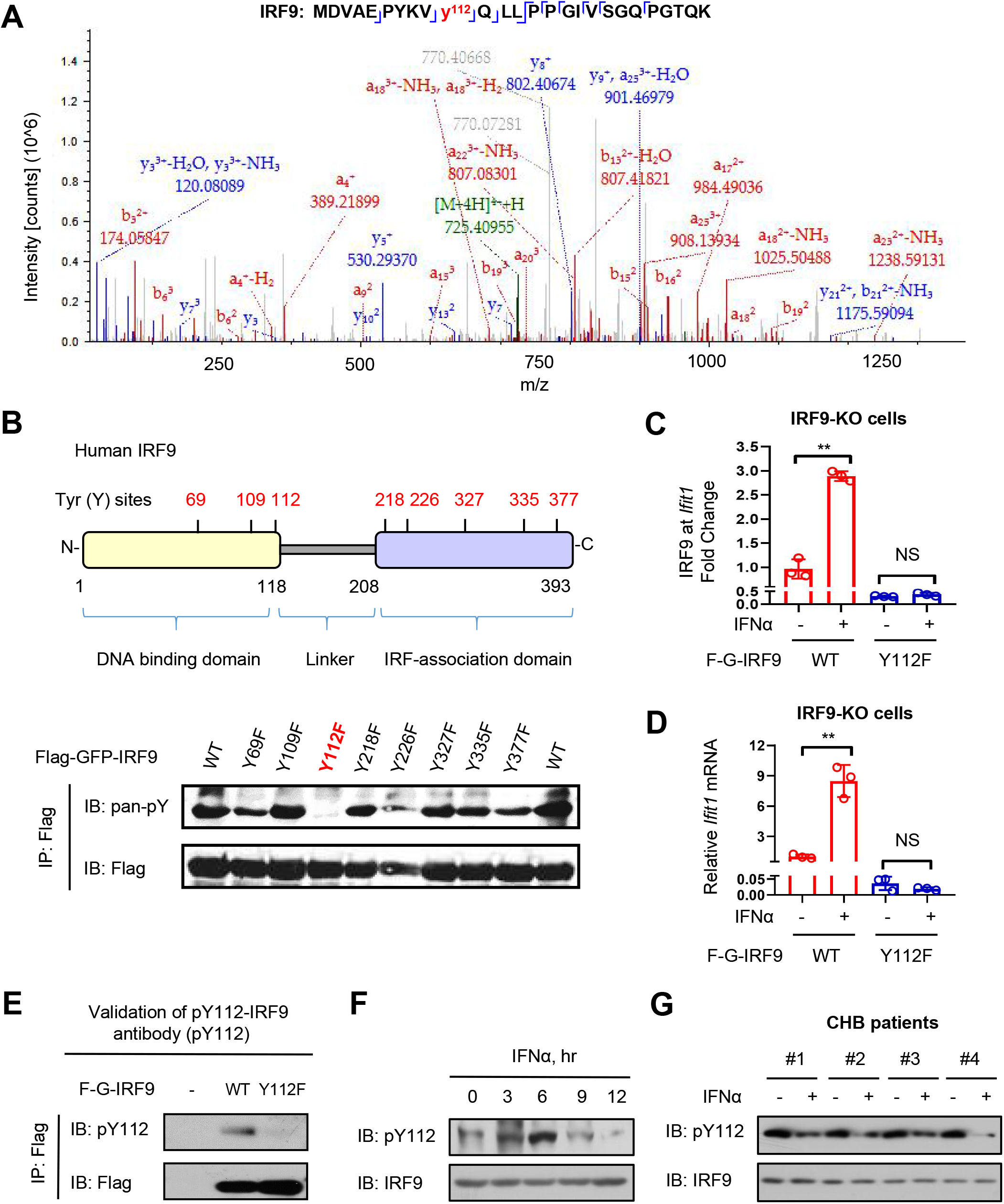
IRF9 Tyr112 phosphorylation is critical for IRF9-ISRE binding but is reduced with durable IFN-I treatment. (A) The tyrosine phosphorylation site (Tyr112) of IRF9 was identified by mass spectrometry. (B) All tyrosine residues of IRF9 were shown (upper). Immunoprecipitation (IP)-immunoblotting (IB) was performed to analyze pan-tyrosine phosphorylation (pan-pY) of IRF9 in HEK293T cells transfected with WT or mutated Flag-GFP-tagged IRF9 (F-G-IRF9) (lower). (C and D) ChIP-qPCR analysis of the binding of IRF9 proteins to the ISRE of *Ifit1* (C) or RT-qPCR analysis of *Ifit1* mRNA levels (D) in IRF9-KO cells stably transfected with F-G-IRF9 (WT, Y112F) and then treated with IFNα (1,000 IU/ml) for 8 h. (E) IP-IB analysis of pY112-IRF9 using a specific anti-pY112-IRF9 antibody in HEK293T cells transfected with F-G-IRF9 (WT or Y112F). (F) Western blot analysis of pY112-IRF9 in HEK293T cells treated with IFNα (1,000 IU/ml) for indicated times. (G) Western blot analysis of pY112-IRF9 in PBMCs from the CHB patients in Figure 1B. B: before Peg-IFN treatment; A: 6 h after Peg-IFN treatment. NS, not significant (p > 0.05), **p < 0.01 (two-tailed unpaired Student’s *t*-test). Data are shown as means ± SD of three biological replicates (C, D), or are representative of three independent experiments (B, E-G).

Next, we made a specific antibody against pY112-IRF9 (Fig. 2E and S1E). We confirmed that this specific pY112-IRF9 antibody can recognize only wild-type IRF9 but not Tyr112-mutated IRF9 (Fig. 2E). We noticed that IRF9 proteins in various types of cells undergo Tyr112 phosphorylation modifications, which were upregulated at the early stage of IFN-I treatment and then were gradually lowered with durable IFN-I treatment (Fig. 2F and S1F). And the timing downregulating pY112-IRF9 levels and attenuating IRF9-ISRE binding was consistent (Fig. 2F and 1E). In line with these results, the pY112-IRF9 levels in PBMCs of chronic CHB patients were reduced after Peg-IFN treatment (Fig. 2G). Collectively, these findings demonstrated that IRF9 Tyr112 phosphorylation is critical for IRF9-ISRE binding and subsequent ISG transcriptional expression, while prolonged IFN-I stimulation leads to pY112-IRF9 de-phosphorylation, thus restricting the durability of IFN-I signaling.

### Salicylate maintains pY112-IRF9 and promotes IRF9-ISRE binding for ISGs expression during IFN-I signaling

Given that pY112-IRF9 levels are critical for IFN-I signaling durability, we sought to explore the potential drugs to improve the level of pY112-IRF9 during IFN-I signaling. To this end, a drug library containing 214 small molecules from plant sources, which have been approved for clinical treatment, was employed. Interestingly, we noticed that both Aspirin (acetylsalicylic acid) and its metabolite in the body, Salicylic acid, could strongly elevate pY112-IRF9 levels in cells with IFN-I treatment (Fig. 3A and S2). Further studies confirmed that both Aspirin and Salicylate (a salt of Salicylic acid) can substantially block pY112-IRF9 reduction induced by IFN-I (Fig. 3B and S3A). Since Aspirin is metabolized *in vivo* mainly as Salicylate and then distributed throughout various tissues of the whole body, we next focused on Salicylate to study its effects on IFN-I signaling.

**Figure 3.**
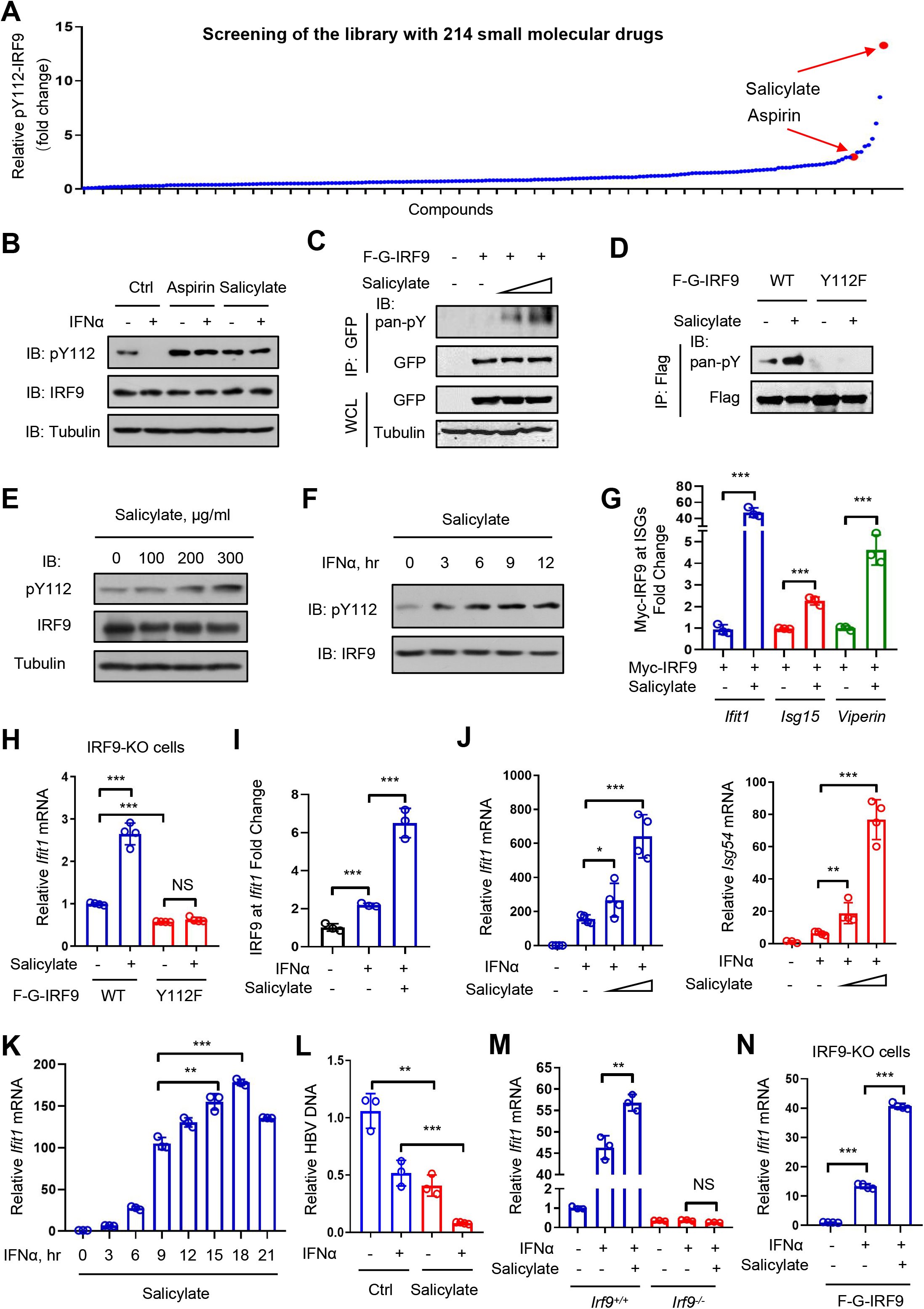
Salicylate maintains pY112-IRF9 and promotes IRF9-ISRE binding for ISGs expression during IFN-I signaling. (A) A drug library containing 214 clinically approved small molecules from plant sources was employed to screen for the potential small molecules that can upregulate pY112-IRF9 in HEK293T cells with IFNα treatment (1,000 IU/ml, 12 h). (B) Western blot analysis of pY112-IRF9 in HEK293T cells pre-treated with 1×PBS (Ctrl) or Aspirin (100 μg/ml) or Salicylate (100 μg/ml) for 6 h and then treated with or without IFNα (1,000 IU/ml) for 12 h. (C) IP-IB analysis of pan-pY of IRF9 in HEK293T cells transfected with F-G-IRF9 and treated with Salicylate (300 and 600 μg/ml) for 6 h. (D) IP-IB analysis of pan-pY of IRF9 in HEK293T cells transfected with F-G-IRF9 (WT, Y112F) and treated with Salicylate (100 μg/ml) for 6 h. (E and F) Western blot analysis of pY112-IRF9 in HEK293T cells treated with Salicylate (100, 200 and 300 μg/ml) for 6 h (E) or Salicylate (100 μg/ml) for indicated times (F). (G) ChIP-qPCR analysis of the binding of IRF9 to the ISRE of *Ifit1*, *Isg15* or *Viperin* genes in HEK293T cells transfected with Myc-IRF9 and treated with Salicylate (300 μg/ml) for 6 h. (H) RT-qPCR analysis of *Ifit1* mRNA in IRF9-knockout (KO) cells stably transfected with F-G-IRF9 (WT, Y112F) and treated with Salicylate (300 μg/ml) for 6 h. (I) ChIP-qPCR analysis of the binding of IRF9 to the ISRE of *Ifit1* in 2fTGH cells pre-treated with Salicylate (300 μg/ml) for 2 h and then treated with IFNα (1,000 IU/ml) for 6 h. (J) RT-qPCR analysis of *Ifit1* and *Isg54* mRNA in THP1 cells pre-treated with Salicylate (100 and 300 μg/ml) for 2 h and then treated with IFNα (1,000 IU/ml) for 6 h. (K) RT-qPCR analysis of *Ifit1* mRNA in HEK293T cells treated with Salicylate (100 μg/ml) and IFNα (1,000 IU/ml) together for indicated times. (L) HepG2 cells transfected with HBV-1.3 constructs were treated with Salicylate (300 μg/ml) and (or) IFNα (1,000 IU/ml) together for 24 h. RT-qPCR was used to analyze HBV DNA levels in cells. (M) RT-qPCR analysis of *Ifit1* mRNA in *Irf9^+/+^* and *Irf9^-/-^* cells pre-treated with Salicylate (300 μg/ml, 2 h) and then treated with IFNα (1,000 IU/ml) for 6 h. (N) RT-qPCR analysis of *Ifit1* mRNA in *Irf9^-/-^* HEK293T cells transfected with F-G-IRF9 and treated with Salicylate (100 μg/ml, 2 h), followed by IFNα treatment (1,000 IU/ml) for 6 h. NS, not significant (p > 0.05), *p < 0.05, **p < 0.01, ***p < 0.001 (two-tailed unpaired Student’s *t*-test). Data are shown as means ± SD of at least three biological replicates (G-N), or are representative of three independent experiments (B-F).

We found that Salicylate upregulated IRF9 tyrosine phosphorylation (Fig. 3C and S3B), but did not affect IRF9 acetylation (Fig. S3C). Furthermore, mutation of Tyr112 on IRF9 abolished Salicylate-mediated upregulation of pY112-IRF9 (Fig. 3D). Using the specific antibody against pY112-IRF9, we demonstrated that Salicylate upregulated pY112-IRF9 levels in a dose- and time-dependent manner (Fig. 3E and S3D). Importantly, Salicylate blocked downregulation of pY112-IRF9 during IFN-I signaling (Fig. 3F). In line with pY112-IRF9 upregulation, Salicylate significantly promoted IRF9 binding to the ISRE of ISGs (Fig. 3G and S3E), and improved ISG transcription expression in cells with wild-type IRF9 but not Tyr112-mutated IRF9 (Fig. 3H). Furthermore, we observed the effects of Salicylate on IFN-I-induced IRF9-ISRE binding and ISG transcription. The results showed that Salicylate enhanced IFN-I-stimulated IRF9-ISRE binding (Fig. 3I) and the activity of ISRE promoters (Fig. S3F). Consistently, Salicylate improved IFN-I-induced ISG transcriptional expression in a dose-dependent (Fig. 3J and S3G) and time-dependent (Fig. S4A) manner. Similarly, Salicylate-mediated enhancement of ISG mRNA and protein expression can be observed in various types of cells (Fig. S4B-J). This is not due to higher IFN-I production, since Salicylate did not promote IFN-I production during virus infection (Fig. S4K). Importantly, we found that Salicylate treatment substantially delayed the decay of IFN-I signaling (Fig. 3K).

We further found that Salicylate promoted IFN-I-mediated inhibition of HBV infection in liver cells (Fig. 3L). In addition, given that IFN-I exerts broad-spectrum antiviral activity, we observed the effect of Salicylate on IFN-I antiviral activity using other model viruses. The results showed that Salicylate also enhanced IFN-I-mediated inhibition of infection of Vesicular Stomatitis Virus (VSV) (Fig. S5A) and influenza A virus (H1N1) (Fig. S5B). Consistently, Salicylate treatment inhibited infection of various types of viruses in cells (Fig. S5C). Salicylate-mediated inhibition of virus infection was also observed in many different cell types (Fig. S5D and S5E), and was dose-dependent (Fig. S5F-H). We further demonstrated that the effects of Salicylate on IFN-I signaling are dependent of IRF9, since IRF9 knockout abolished Salicylate-mediated enhancement of ISG transcriptional expression (Fig. 3M and S6A). Reconstitution of IRF9 in *Irf9^-/-^* cells rescued Salicylate-mediated enhancement of IFN-induced ISG expression (Fig. 3N). Consistently, knockout of IFNAR1 but neither STAT1 nor STAT2 abolished Salicylate-mediated ISG upregulation (Fig. S6B-E). Moreover, mutation of Tyr112 on IRF9 blocked Salicylate-mediated inhibition of virus infection (Fig. S6F). Taken all together, these findings demonstrated that Salicylate promotes IRF9-Tyr112 phosphorylation, IRF9-ISRE binding, ISG transcriptional expression and IFN-I antiviral activity in cells with IFN-I treatment.

### Salicylate inhibits PTP1B-mediated IRF9 de-phosphorylation at Tyr112

To explore how Salicylate upregulates pY112-IRF9, we first determined whether Salicylate affects activation of STAT1 and STAT2 upstream of IRF9. We noticed that Salicylate did not promote IFN-I-induced activation of either STAT1 (Fig. 4A and S7A) or STAT2 (Fig. S7B). Interestingly, Aspirin remarkably promoted IFN-I-induced STAT1 activation (Fig. 4B), suggesting that Aspirin could have additional regulatory effects on IFN-I signaling. Given that Salicylate did not promote STAT1 and STAT2 activation, we speculated that Salicylate could directly affect the process of IRF9-Tyr112 phosphorylation. Thus, we next utilized different types of the phosphatase inhibitor (PPi) cocktail to observe whether the phosphatases are involved in Salicylate-mediated regulation of IRF9-Tyr112 phosphorylation. The results showed that inhibition of the protein tyrosine phosphatase (PTP) family abolished Salicylate-mediated upregulation of pY112-IRF9 (Fig. 4C and 4D). Consistent with the observation of the phosphatases, our analysis of IRF9-binding proteins by mass spectrometry showed that the phosphatase PTP1B could interact with IRF9 (Fig. 4E). Immunoprecipitation analysis confirmed that PTP1B interacts with both endogenous IRF9 (Fig. 4F) and exogenously expressed F-G-IRF9 (Fig. 4G and S7C). Importantly, PTP1B overexpression downregulated pY-IRF9 levels in a dose-dependent manner (Fig. 4H), while PTP1B knockdown upregulated pY-IRF9 levels (Fig. S7D). However, PTP1B cannot decrease tyrosine phosphorylation of Tyr112-muated IRF9 (Fig. 4I). These data suggested that PTP1B mediates pY112-IRF9 de-phosphorylation.

**Figure 4.**
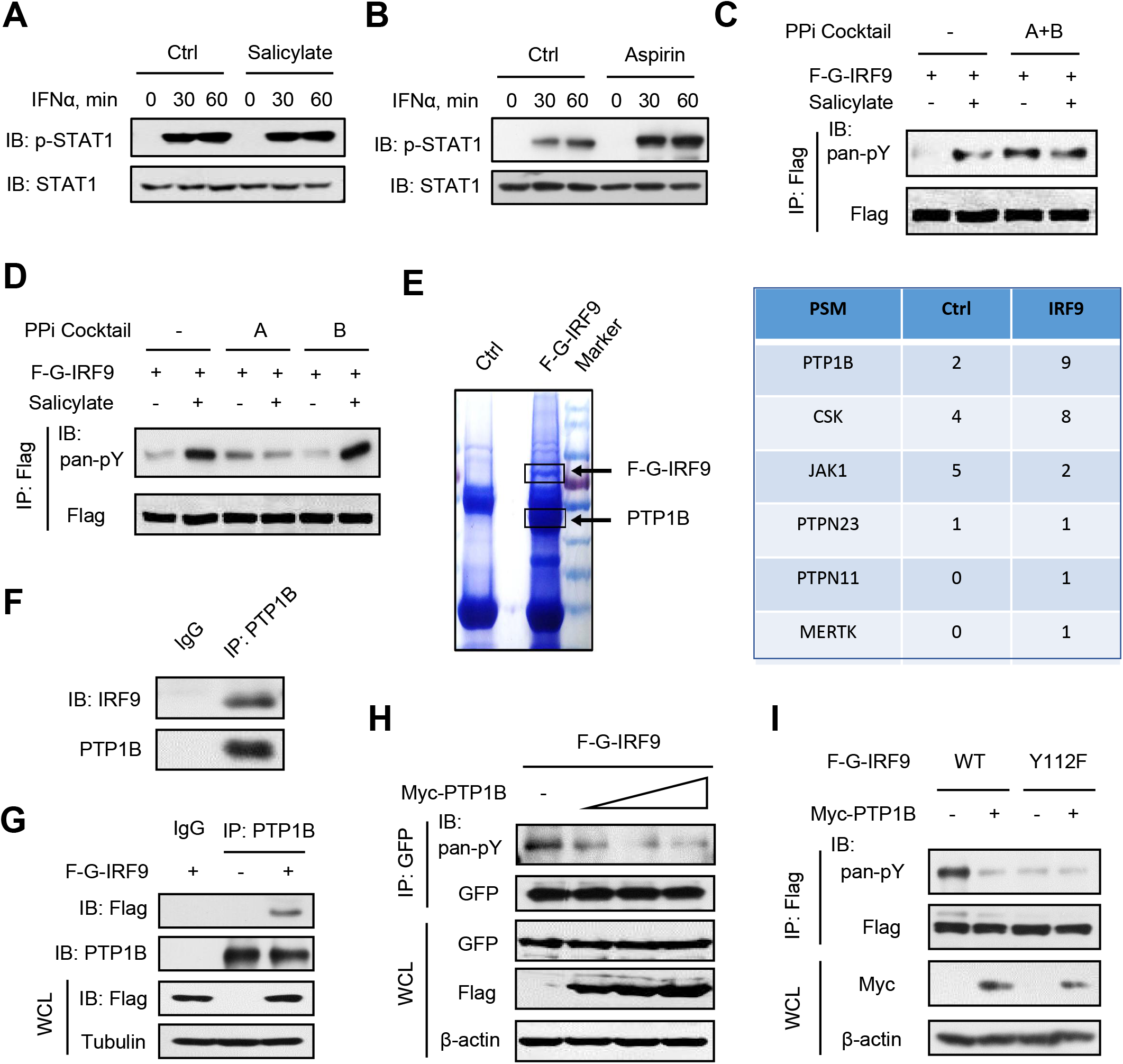
Salicylate inhibits PTP1B-mediated IRF9 de-phosphorylation at Tyr112. (A and B) Western blot analysis of pY701-STAT1 in HEK293T cells pre-treated with Salicylate (100 μg/ml) (A) or Aspirin (100 μg/ml) (B) for 6 h and then treated with IFNα (1,000 IU/ml) as indicated. (C and D) 2fTGH cells transfected with F-G-IRF9 were pre-treated with phosphatase inhibitors (PPi) Cocktail A+B (1:1,000) (C) or PPi Cocktail A or B (1:1,000) (D) for 2 h. Then cells were treated with or without Salicylate (300 μg/ml) for 6 h. IP-IB was performed to analyze pan-pY of IRF9. (E) F-G-IRF9 proteins were co-immunoprecipitated by anti-Flag antibodies from HEK293T cells transfected with F-G-IRF9 (left). Mass spectrometry was used to analyze the potential IRF9-interacting proteins, among which the tyrosine kinases and phosphatases were shown (right). (F) IP-IB analysis of the interaction between the endogenous IRF9 and PTP1B in HEK293T cells. (G) IP-IB analysis of the interaction between F-G-IRF9 and endogenous PTP1B in HEK293T cells transfected with F-G-IRF9. (H) IP-IB analysis of pan-pY of F-G-IRF9 in HEK293T cells transfected with F-G-IRF9 and increased amounts of Myc-PTP1B. (I) IP-IB analysis of pan-pY of F-G-IRF9 in HEK293T cells transfected with F-G-IRF9 (WT, Y112F), together with or without Myc-PTP1B. Data are representative of three independent experiments (A-D, F-I).

We observed that Salicylate treatment inhibited the interaction between PTP1B and IRF9 in a time-dependent and dose-dependent manner (Fig. 5A, S7E and S7F). Similarly, Salicylate inhibited PTP1B binding with exogenously expressed F-G-IRF9 (Fig. 5B and S7G). In addition, IFN-I treatment can enhance PTP1B-IRF9 interaction, which was inhibited by Salicylate (Fig. 5C). Consistently, Salicylate treatment inhibited PTP1B-mediated de-phosphorylation of pY-IRF9 (Fig. 5D). Moreover, we demonstrated that Salicylate treatment blocked PTP1B-mediated de-phosphorylation at Tyr112 of IRF9 (Fig. 5E). Taken together, these findings suggested that Salicylate could inhibit PTP1B-IRF9 interaction to maintain pY112-IRF9 levels in IFN-I signaling. Next, in order to observe the possible interaction between Salicylate and PTP1B, we employed a NanoDSF assay. The results clearly supported Salicylate-PTP1B interaction, as shown in the observation that Salicylate improved the melting temperature (Tm) value by 1.38 ℃ (Fig. 5F), suggesting that it greatly improved PTP1B thermo-stability. Furthermore, the sodium Salicylate was docked onto the PTP1B protein using AutoDock Vina (Trott and Olson, 2010), and the top three ranking poses of each docked ligand were observed (Fig. 5G). The results showed that the top position is at PTP1B active pocket with a calculated Gibbs free energy (ΔG) of -5.851 Kcal/mol, while the other two positions are located in the vicinity of active pocket and the backside of PTP1B (Fig. 5G), suggesting that Salicylate preferentially binds to the PTP1B active pocket. In addition, the interactions between PTP1B and Salicylate include two categories. One is hydrophilic interactions, which mainly includes carboxylate group of Salicylate: it forms salt bond or hydrogen bonds with surrounding residues S216, A217, G218, and R221 (Fig. 5H). The other is hydrophobic interactions, which mainly includes aromatic ring group of Salicylate: it interacts with side chains of F182, I219 and R221 (Fig. 5H). All these interactions form the network to stabilize Salicylate in PTP1B active pocket (Fig. 5I). Taken all together, these observations suggested that Salicylate could interact with PTP1B and restrict PTP1B-mediated de-phosphorylation of IRF9.

**Figure 5.**
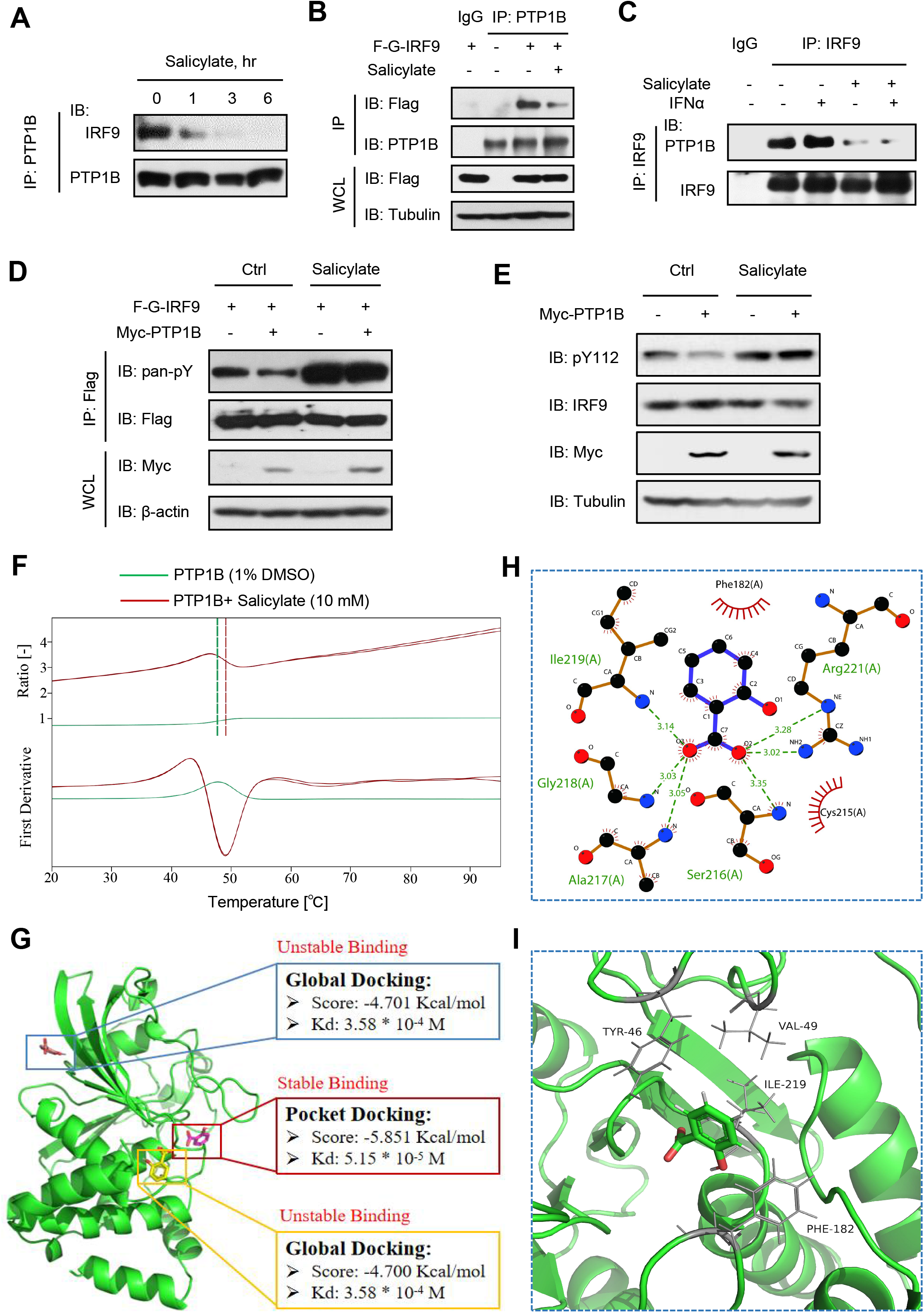
Salicylate inhibits the interaction between PTP1B and IRF9. (A) IP-IB analysis of the interaction between IRF9 and PTP1B in HEK293T cells treated with Salicylate (300 μg/ml) for indicated times. (B) IP-IB analysis of the interaction between F-G-IRF9 and PTP1B in HEK293T cells transfected with F-G-IRF9 and treated with Salicylate (300 μg/ml) for 6 h. (C) IP-IB analysis of the IRF9-PTP1B interaction in HEK293T cells pre-treated with Salicylate (300 μg/ml) for 2 h and then treated with IFNα (1,000 IU/ml) for 6 h. (D) IP-IB analysis of pan-pY of IRF9 in HEK293T cells transfected with F-G-IRF9 and Myc-PTP1B, and then treated with or without Salicylate (300 μg/ml) for 6 h. (E) Western blot analysis of pY112-IRF9 in HEK293T cells transfected with Myc-PTP1B, and then treated with or without Salicylate (100 μg/ml) for 6 h. (F) The nanoDSF study of the effect of Salicylate on PTP1B thermal stability. The upper panel shows the fluorescence signal ratio change between 350 nm and 330 nm. The lower panel shows the derivative values. Vertical dashed lines show the melting temperatures. (G) Docking of Salicylate to PTP1B. PTP1B is shown as a cartoon and Salicylates are shown as sticks. The calculated binding energies and the equilibrium dissociation constants (Kd) are shown in the rectangle boxes. (H) Predicted hydrophilic interactions between Salicylate and PTP1B at its active pocket. All atoms are shown as balls and sticks, where carbon atoms are colored black, oxygen atoms are red and nitrogen atoms are blue. Chemical bonds are shown as blue or brown sticks. Predicted hydrogen bonds between Salicylate and PTP1B are shown as dashed lines in angstrom. (I) Predicted hydrophobic interactions between Salicylate and PTP1B at its active pocket. Salicylate is show as sticks and PTP1B residues interacting with Salicylate are shown as thin sticks including hydrogen atoms. Data are representative of three independent experiments (A-E).

### Aspirin improves the reactivity of IFN-I signaling by attenuating PARP11-β-TrCP axis to stabilize IFNAR1

Our aforementioned results suggested that Aspirin can promote IFN-I-induced STAT1 activation (Fig. 4B), which can be further confirmed in cells with IFN-I treatment for a longer period (Fig. 6A). Here, we further explored whether Aspirin has additional activities to promote IFN-I signaling. Given that Aspirin enhanced IFN-I-induced STAT1 phosphorylation, we speculated that Aspirin could have a target upstream of STAT1 activation. Thus, we first determined whether Aspirin treatment could affect IFN-I receptors (IFNAR1 and IFNAR2). We noticed that in cells with Aspirin treatment, protein levels of IFNAR1 but not IFNAR2 were remarkably upregulated in a time-dependent manner (Fig. 6B). However, Aspirin did not upregulate IFNAR1 mRNA levels (Fig. 6C). We further found that Aspirin but not Salicylate largely blocked IFN-I-induced IFNAR1 downregulation (Fig. 6D and 6E), suggesting that Aspirin facilitates IFNAR1 protein stability. Consistently, Aspirin treatment attenuated IFNAR1 ubiquitination during IFN-I signaling (Fig. 6F). These findings suggested that Aspirin is able to inhibit IFNAR1 degradation in IFN-I signaling.

**Figure 6.**
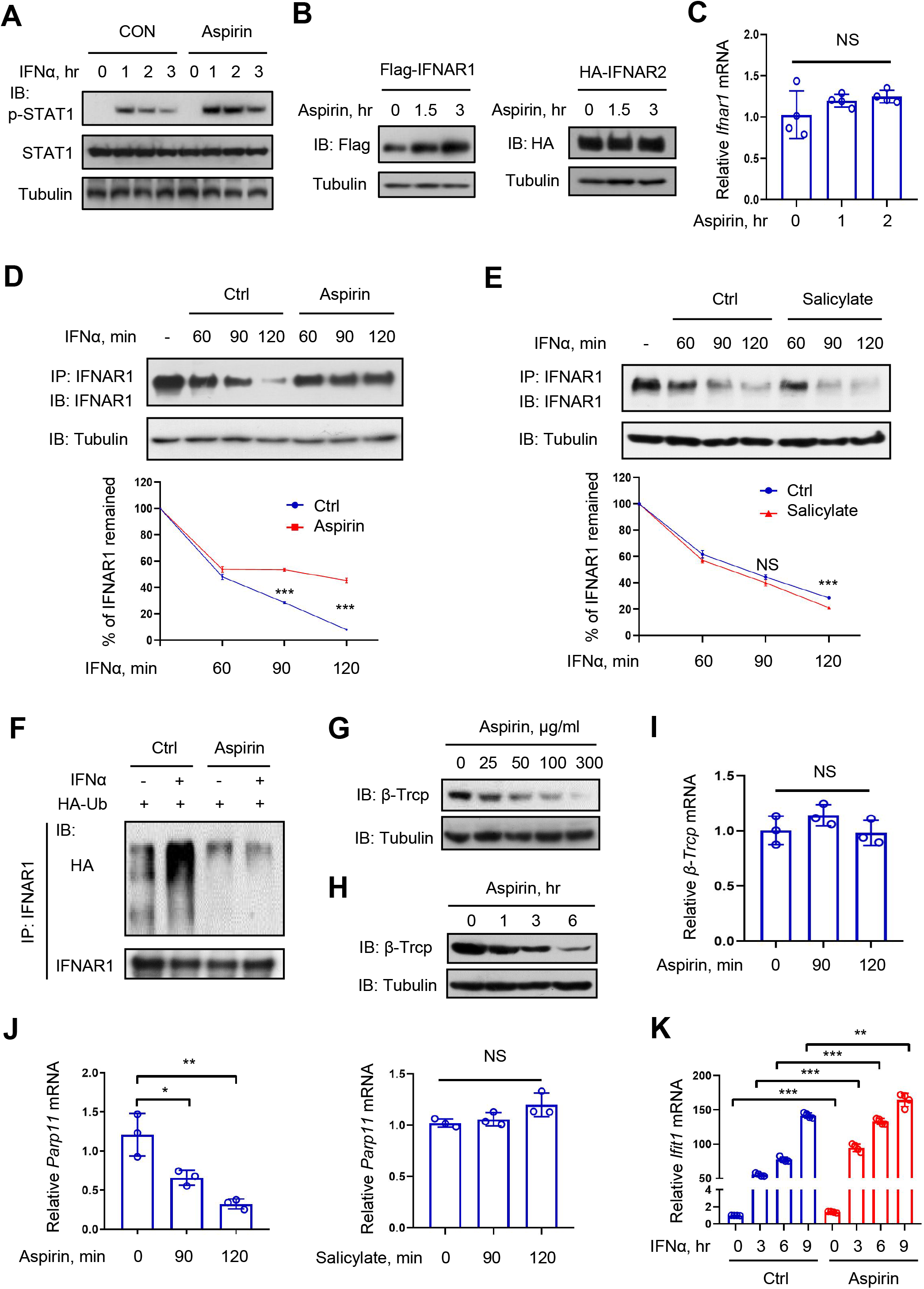
Aspirin improves the reactivity of IFN-I signaling by attenuating PARP11-β-TrCP axis to stabilize IFNAR1. (A) Western blot analysis of pY701-STAT1 in HEK293T cells treated with Aspirin (100 μg/ml) and (or) IFNα (1,000 IU/ml) as indicated. (B) Western blot analysis of Flag-IFNAR1 or HA-IFNAR2 in HEK293T cells transfected with either Flag-IFNAR1 (left) or HA-IFNAR2 (right) and then treated with Aspirin (100 μg/ml) as indicated. (C) RT-qPCR analysis of *Ifnar1* mRNA in HEK293T cells treated with Aspirin (100 μg/ml) for indicated times. (D and E) Western blot analysis of IFNAR1 in HEK293T cells pre-treated with Aspirin (100 μg/ml) (D) or Salicylate (100 μg/ml) (E) for 2 h and then treated with IFNα (2,000 IU/ml) for 60, 90 and 120 min. (F) HEK293T cells transfected with HA-tagged ubiquitin (Ub) were pre-treated with Aspirin (100 μg/ml, 2 h) and then treated with IFNα (2,000 IU/ml) for 2 h. IP-IB was performed to analyze IFNAR1 ubiquitination in cells. (G and H) Western blot analysis of β-TrCP in HEK293T cells treated with Aspirin for 6 h as indicated (G) or Aspirin (100 μg/ml) for indicated times (H). (I) RT-qPCR analysis of *β-TrCP* mRNA in HEK293T cells treated with Aspirin (100 μg/ml) for indicated times. (J) RT-qPCR analysis of *Parp11* mRNA in HEK293T cells treated with Aspirin (100 μg/ml) (left) or Salicylate (100 μg/ml) (right) for indicated times. (K) RT-qPCR analysis of *Ifit1* mRNA in HEK293T cells pre-treated with Aspirin (100 μg/ml) for 2 h and then treated with IFNα (1,000 IU/ml) as indicated. NS, not significant (p > 0.05), *p < 0.05, **p < 0.01, ***p < 0.001 (two-tailed unpaired Student’s *t*-test). Data are shown as means ± SD of at least three biological replicates (C, I-K), or are representative of three independent experiments (A, B, D-H).

IFNAR1 protein stability has been proven to be critically regulated by the ubiquitin E3 ligase β-TrCP (Kumar et al., 2004; Kumar et al., 2003). We further found that Aspirin treatment can reduce β-TrCP protein levels in both a dose-dependent (Fig. 6G) and time-dependent (Fig. 6H) manner. However, Aspirin treatment did not significantly affect β-TrCP mRNA levels (Fig. 6I). One of my recent studies revealed that β-TrCP ADP-ribosylation mediated by ADP-ribosyltransferase PARP11 protects β-TrCP from ubiquitin-proteasome degradation and promotes IFNAR1 ubiquitination and degradation (Guo et al., 2019). Thus, we observed the possible effect of Aspirin or Salicylate on PARP11. Interestingly, we found that Aspirin but not Salicylate significantly reduced PARP11 mRNA levels in a time-dependent manner (Fig. 6J). Consistent with the attenuated PARP11-β-TrCP axis, Aspirin significantly improved the strength of IFN-I signaling (Fig. 6K). Although the detailed mechanism by which Aspirin regulates PARP11 remains to be elucidated, our studies here clearly demonstrated that Aspirin is able to reduce cellular PARP11 and β-TrCP levels, thus facilitating IFNAR1 protein stability and in turn enhancing IFN-I signaling reactivity.

### Aspirin improves both the reactivity and durability of IFN-I signaling *in vivo*

Our findings above demonstrated that both Salicylate and Aspirin in cells can maintain pY112-IRF9 levels and Aspirin can maintain IFNAR1 stability during IFN-I signaling. Given that Aspirin is metabolized *in vivo* mainly as Salicylate that is distributed throughout the whole body, we further explored whether Aspirin administration benefits *in vivo* IFN-I signaling. To this end, mice were injected intraperitoneally (*i.p.*) with or without Aspirin (50 μg per gram body mouse). Two hours after Aspirin administration, mice were treated with mouse IFN-I (mIFNβ, *i.p.*) for 6 h (Fig. 7A). We noticed that IFN-I activated ISG expression in mouse spleens, while Aspirin administration significantly promoted IFN-I-induced expression of several representative ISGs (Fig. 7B). Similarly, Aspirin administration substantially improved IFN-I-induced ISG expression in various tissues of mice, including mouse heart, liver, lung and kidney tissues (Fig. 7C). Furthermore, we found that pY112-IRF9 levels in mouse lung tissues were reduced at 6 h after mIFNβ treatment, while Aspirin administration maintained pY112-IRF9 levels (Fig. 7D). Moreover, we found that IFNAR1 protein levels were downregulated after IFN-I treatment, while Aspirin largely blocked IFNAR1 downregulation (Fig. 7E). Consistent with the enhancement of *in vivo* IFN-I signaling, Salicylate administration significantly inhibited virus infection in various tissues of mice (Fig. S8A-E).

**Figure 7.**
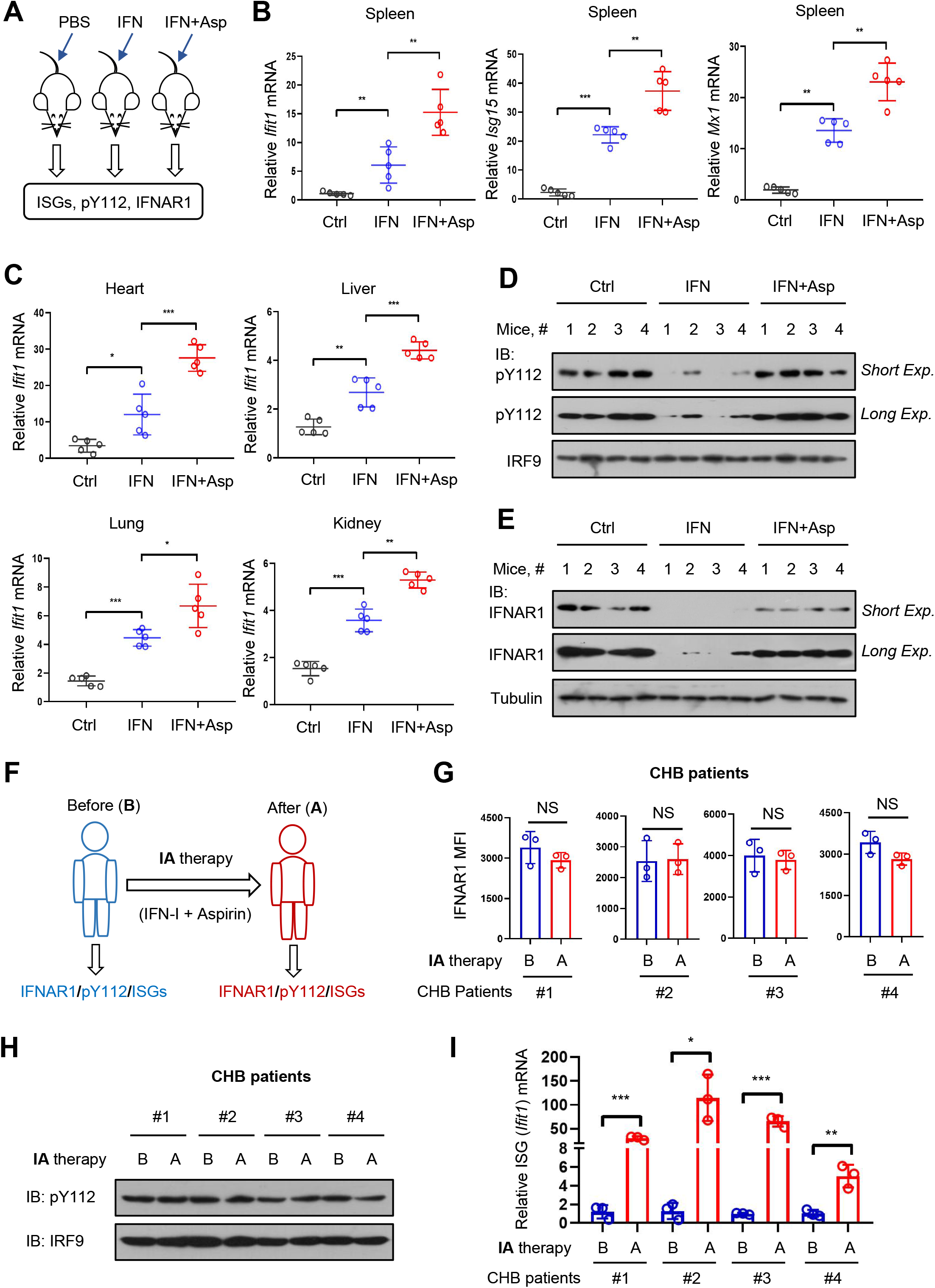
Aspirin improves both the reactivity and durability of IFN-I signaling *in vivo*. (A) Mice (n = 5) were injected intraperitoneally (*i.p.*) with PBS or Aspirin (50 μg per gram body mouse). Two hours after injection, mice were injected with mouse IFNβ (1,000 IU/g, *i.p.*) and then mouse tissues were collected for further analysis 6 h after IFNβ injection. (B) RT-qPCR analysis of *Ifit1*, *Isg15* and *Mx1* mRNA in mouse spleen tissues from (A). (C) RT-qPCR analysis of *Ifit1* mRNA in mouse heart, liver, lung and kidney tissues from (A). (D and E) Western blot analysis of pY112-IRF9 (D) and IFNAR1 (E) in the livers of mice (n=4) injected with Aspirin and IFNβ as (A). (F) CHB patients were recruited and the IA (IFN-I + Aspirin) therapy was performed. Aspirin (0.2 g, qw) was taken orally before Peg-IFN injection and then Peg-IFN (180 μg, qw) was injected subcutaneously in the abdomen. The PBMCs from CHB patients were collected for analysis before injection and 6 h after injection. B: before injection. A: 6 h after injection. (G) Flow cytometry analysis of IFNAR1 in PBMCs of CHB patients from (F). (H) Western blot analysis of pY112-IRF9 in PBMCs of CHB patients from (F). (I) RT-qPCR analysis of *Ifit1* mRNA in PBMCs of CHB patients from (F). NS, not significant (p > 0.05), *p < 0.05, **p < 0.01, ***p < 0.001 (two-tailed unpaired Student’s *t*-test). All graphs show the mean ± SEM for five individual mice (B, C). Data are shown as means ± SD of three biological replicates (G, I), or are representative of three independent experiments (D, E, H).

Aspirin has been widely used in clinical therapy and has the potential to reduce fever caused by IFN-I treatment. Thus, we recruited 18 patients with chronic HBV infection, who were diagnosed to be suitable for IFN-I treatment (Table 1). These patients took Aspirin (0.2 g, once a week) orally before the weekly IFN-I injection. The combination therapy of IFN-I and Aspirin was named IA therapy. Then we collected the PBMCs from the first four recruited patients at 6 h after the first IFN-I injection (Fig. 7F). We noticed that IFNAR1 protein levels in the PBMCs from these patients with IA therapy remained relatively stable after IFN-I treatment (Fig. 7G). In addition, downregulation of pY112-IRF9 after IFN-I treatment was largely inhibited in these patients with IA therapy (Fig. 7H). Importantly, we observed that IA therapy resulted in much higher expression levels of ISGs in these patients (Fig. 7I), which shows more efficient IFN-I signaling as compared to IFN-I-only therapy (Fig. 1C). Collectively, these observations suggested that Aspirin could substantially advance *in vivo* IFN-I signaling by stabilizing IFNAR1 and maintaining pY112-IRF9 levels.

**Table 1.** Information of CHB patients before and after IA therapy.

| | IFNα + Aspirin |
| --- | --- |
| Patients enrolled, n | 15 |
| Age, years <sup>a</sup> | 42.27±6.61 |
| Male, n (%) | 12 (80%) |
| HBsAg (baseline), IU/ml | 0-1500 |
| 500 < HBsAg ≤ 1500 IU/ml, n (%) | 3 (20%) |
| 10 < HBsAg ≤ 500 IU/ml, n (%) | 6 (40%) |
| 0 < HBsAg ≤ 10 IU/ml, n (%) | 6 (40%) |
| HBsAg loss (48 weeks), n (%) | 13 (86.67%) |
| 500 < HBsAg ≤ 1500 IU/ml, n (%) | 2 (66.67%) |
| 10 < HBsAg ≤ 500 IU/ml, n (%) | 5 (83.33%) |
| 0 < HBsAg ≤ 10 IU/ml, n (%) | 6 (100%) |
| HBV DNA undetectable (48<sup>th</sup> week), n (%) | 15 (100%) |
| No fever (during 1<sup>st</sup> therapy), n (%) | 11 (73.33%) |

<sup>a</sup>The data conforms to a normal distribution and is shown as means ± SD.

### Aspirin remolds the therapy for a functional cure of chronic HBV infection

We continued to implement IA therapy and observation for all recruited patients. During 48 weeks of IA therapy, three patients left our observation, either forgetting to take Aspirin or losing contact. Finally, 15 patients in total completed the IA therapy (Fig. 8A). The data of HBsAg loss and HBV DNA detection were shown in the Table 1. Consistent with the observation of substantially improved IFN-I signaling in the patients (Fig. 7G-I), 13 out of 15 CHB patients with IA therapy achieved a functional cure within 48 weeks. In detail, 2 out of 3 patients with initial HBsAg levels between 500-1500 IU/ml achieved a functional cure within 48 weeks, and the third patient with HBsAg between 500-1500 IU/ml achieved a functional cure within 72 weeks of IA therapy (Fig. 8B). 5 out of 6 CHB patients with initial HBsAg levels between 10-500 IU/ml achieved a functional cure within 48 weeks, and among these five patients, four patients got this end-point within 28 weeks (Fig. 8C). 6 out of 6 CHB patients with initial HBsAg levels under 10 IU/ml achieved a functional cure within 20 weeks (Fig. 8D). In addition, given that Aspirin has a fever-reducing effect, we paid special attention to the patient’s fever. We noticed that although low dosage of Aspirin was used in this study, only 4 out of 15 (26.7%) patients had a fever during the first IFN-I treatment. This percentage of fever is much lower than the reported one of fever caused by IFN-only therapy in the literature (Iftikhar et al., 2020). Taken all together, these observations suggested that the IA therapy could be an efficient clinical therapeutic way for achieving a functional cure of chronic HBV infection.

**Figure 8.**
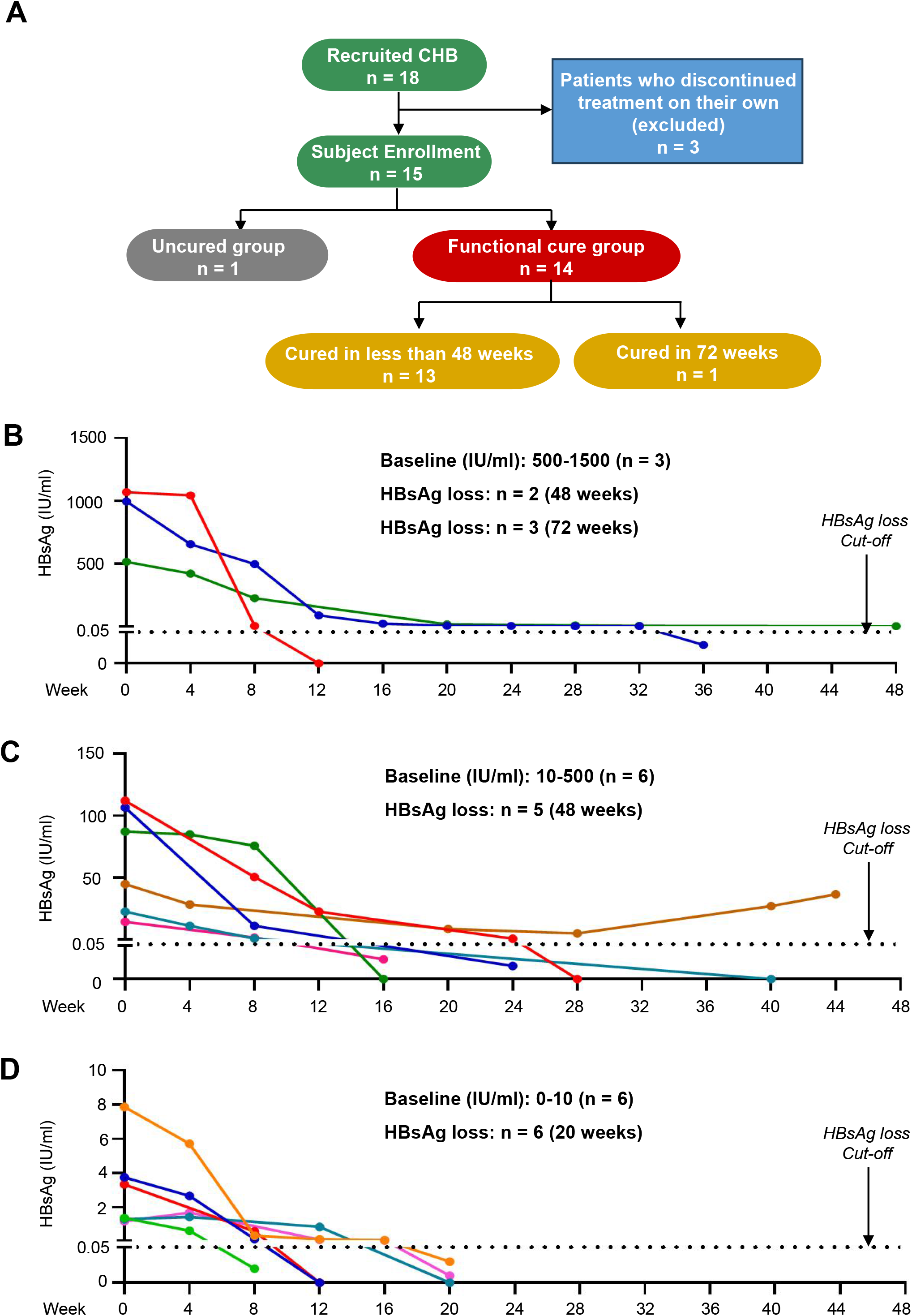
Aspirin remolds the clinical therapy for a functional cure of chronic HBV infection. (A) CHB patients enrolled in the study (n = 18) and the functional cure of CHB patients (n = 15) after the IA therapy. (B-D) CHB Patients were divided into three groups based on the HBsAg levels: 500-1500 IU/ml (B) (n = 3), 10-500 IU/ml (C) (n = 6) and 0-10 IU/ml (D) (n = 6). The changes in HBsAg levels (IU/ml) from baseline to the end-point of IA therapy were shown over time.

## Discussion

In this study, we explored the strategies for improving IFN-I signaling and IFN-I therapeutic efficacy in clinic. During the long-term studies of IFN-I signaling, we and others revealed that IFN-I treatment induces the degradation of its receptor (IFNAR1), thereby inhibiting the subsequent reactivity of IFN-I signaling. Here, we further found that maintaining IFN-I receptor stability can increase only the strength of IFN-I signaling, but still cannot block a rapid decay of IFN-I signaling. As a consequence, the expression of ISGs is rapidly downregulated to a very low level after a period of IFN-I treatment. To delay the IFN-I signaling decay to obtain efficient IFN-I therapeutic activity, we focused on the IRF9-ISRE binding, which indeed determines the activation of ISG transcriptional expression. Our next studies demonstrated that IRF9 phosphorylation at Tyr112 is crucial for IRF9-ISRE binding and ISG expression, while IFN-I treatment induces de-phosphorylation of pY112-IRF9. In line with our idea, inhibition of IRF9 de-phosphorylation significantly delays the IFN-I signaling decay, thus improving the durability of IFN-I signaling. Importantly, this study found Aspirin as an efficient “fuel” *in vivo* to both stabilize IFN-I receptors and maintain IRF9 tyrosine phosphorylation, thus improving both the reactivity and durability of IFN-I signaling (Fig. 9).

**Figure 9.**
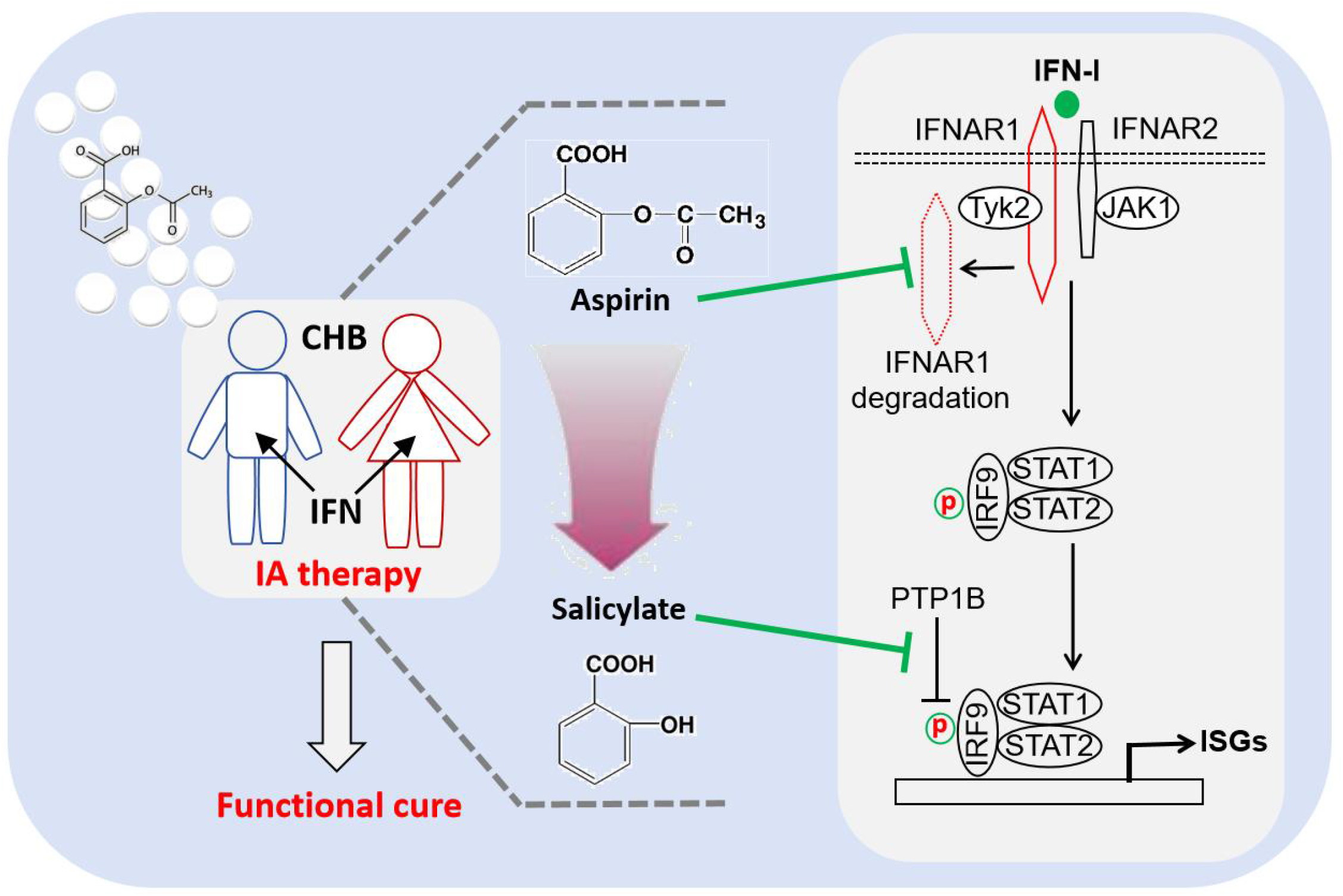
A proposed model. Aspirin and its metabolite Salicylate maintain IFNAR1 protein stability and IRF9 tyrosine phosphorylation, respectively, thus enhancing both the reactivity and durability of IFN-I signaling, which eventually promotes a functional cure of chronic HBV infection.

IFN-I is a cytokine (also ligand) that is widely used for the therapy of hepatitis virus infections and other diseases. Based on the observations from IFN-I signaling, we speculate that many other cytokines (or ligands) have the similar mechanisms to regulate their signaling in cells. It is not difficult to understand that many cellular negative feedback mechanisms are involved in the inhibition of cytokine-mediated signaling. However, based on the studies on IFN-I signaling, we believe that the degradation of cytokine receptors and the decay of the transcriptional expression of cytokine signaling could be two key mechanisms controlling cellular cytokine signaling: one targets the start point of cytokine signaling, and the other targets the end point of cytokine signaling. Degradation of the cytokine receptors will block further stimulation of cells by cytokines, reducing the reactivity of cytokine signaling. Decay of cytokine-induced transcriptional expression will stop producing their effectors, attenuating the durability of cytokine signaling. Thus, the two mechanisms could widely exist in the regulation of many cytokine signaling, which could maintain the homeostasis of cytokine signaling but eventually lead to the “signaling inability” in clinical therapy. Given that many other cytokines have been used for clinical therapy (such as IL-2, IL-10 and PDGF) or are in development (such as GM-CSF, VEGF, IFNγ) (Qiu et al., 2021), we think that our study on IFN-I signaling could provide a paradigm to study how to improve the therapeutic efficacy of these cytokines in clinic.

Aspirin has been widely known as an effective analgesic, antipyretic, anti-inflammatory, and antithrombotic agent, as Aspirin can inhibit prostaglandin and thromboxane biosynthesis mediated by the cyclo-oxygenase-1 (COX-1) and COX-2 isoenzymes (Roth et al., 1975). In addition, recent studies suggested that Aspirin has the potential to provide benefits for cancer prevention (Elwood et al., 2024) and for the treatment of virus infectious diseases, including influenza virus and SARS-CoV-2 infections (Bianconi et al., 2020; Mazur et al., 2007). It has been reported that Aspirin can block influenza virus propagation mainly by inhibiting NF-κB activity (Mazur et al., 2007). However, a recent study demonstrated that Salicylate-mediated inhibition of flavivirus infection is independent of the NF-κB pathway but may involve p38 kinase activity (Liao et al., 2001). As a matter of fact, our study here revealed that Aspirin can enhance IFN-I signaling, which could provide new insight into the effects of Aspirin on cancer prevention and virus inhibition, since IFN-I signaling plays important roles in inhibiting tumor cell proliferation and fighting against broad-spectrum viruses, as well as regulating other signaling pathways (such as the NF-κB pathway). In spite of many reported activities of Aspirin, how Aspirin regulates immune responses, in particular IFN-induced JAK-STAT immune activity, remains less known. Thus, this study could provide clues for understanding of various biological activities of Aspirin *in vivo*. In addition, we showed here that Aspirin could inhibit the activity of PTP1B, which has been demonstrated to be a good target for the therapy of many diseases (such as diabetes and obesity), and PTP1B is reported to be a checkpoint limiting CAR T-cell antitumor immunity (Wiede et al., 2022). Therefore, this study suggests that Aspirin may be a potential adjuvant for the future therapy of these diseases.

A recent meta-analysis reported the latest situation of HBsAg clearance in inactive HBsAg carriers (IHCs) who underwent Peg-IFN therapy (Song et al., 2021). Their analysis collected the data from a total of 1029 IHCs from 11 studies published between January 2000 and August 2021, which showed that the HBsAg loss rates were 92% for HBsAg <10 IU/mL, 60% for HBsAg <500 IU/mL, and 39% for HBsAg <1000 IU/mL in patients after 48 weeks of Peg-IFN therapy (Song et al., 2021). Here, our study showed significantly improved HBsAg loss rates: 100% (6/6) for CHB patients with initial HBsAg <10 IU/mL, 91.7% (11/12) for HBsAg <500 IU/mL, and 86.7% (13/15) for HBsAg <1500 IU/mL within 48 weeks of IA therapy. However, the data presented in this study does not represent the precise value of the HBV functional cure rates caused by the IA therapy. As a matter of fact, this study could be further improved at least by recruiting more HBV patients and adapting the usage of Aspirin. In the current study, Aspirin was used only once a week, and the dosage was relatively low (0.2 g). Thus, further exploration is needed to determine whether additional intake of Aspirin (such as twice a week, before and after Peg-IFN injection) or higher dosage of Aspirin should be used each week to obtain better HBV functional cure rates. In addition, we do not recommend the combination of Aspirin, nucleos(t)ide analogues and IFN-I, as the interactions between these three drugs are not clear and may lead to an unknown outcome of the final HBV functional cure rate. In summary, an optimized IA therapy improved by the clinical professionals is urgently expected to advance the functional cure rate of HBV infections, which can eventually eliminate HBV infections worldwide as soon as possible.

## Data Availability

All data produced in the present study are available upon reasonable request to the authors

## ACKNOWLEDGMENTS

We thank Dr. Serge Y. Fuchs (University of Pennsylvania, USA), Dr. Chen Wang (China Pharmaceutical University), Dr. Eugene Y. Chinn (Soochow University), Dr. Chunfu Zheng (University of Calgary, Canada), Dr. Chunsheng Dong and Dr. Jianfeng Dai (Soochow University) for important reagents. This work is supported by grants from the National Natural Science Foundation of China (32241009), the National Key R&D Program of China (2023YFA1800200) and Suzhou Science and Technology Plan Project (SKY2022063).

## AUTHOR CONTRIBUTIONS

Y.M., Y.Y., F.H., T.Z., R.Z., Q.Z. and Q.C. performed all the molecular, cellular and biochemistry experiments. Y. M., Y.Y., Y.C., J.L., Z.Zhao, M.L., F.Q., L.Z., C.Z. and H.Z. provided patient samples and performed clinical studies. W.T., W.H., Y.Z. and Z.Zheng helped analyze the data. M.L., F.Q., L.Z., C.Z. and H.Z. discussed the data and manuscript. H.Z. conceived the project. H.Z., Y.M., Y.Y. and Y.C. designed the experiments. H.Z. and Y.M. wrote the manuscript.

## METHODS

### Human studies

The clinical research is a single-arm study to evaluate the functional cure of chronic HBV (CHB) patients who complete treatment. This study followed the Good Clinical Practice guidelines and was registered in the Chinese Clinical Trial Registry (ChiCTR) website, and the registration number is ChiCTR2200055870. The subjects of this study were CHB patients administrated with Aspirin (Bayer S.p.A, 0.2 g, orally, qw) first and then injected with Pegylated interferon α-2b (180 μg, qw) in the Affiliated Infectious Diseases Hospital of Soochow University from March 2022 to March 2024. The functional cure was observed whether the patients achieved sustained negative conversion of HBsAg (HBsAg<0.05 IU/ml), with or without HBsAb, HBV DNA below the lowest detection limit, and normal liver function. There were eighteen patients enrolled in this study, among which three patients discontinued treatment due to relocation or poor compliance. Thus 15 patients were finally included in this study. HBV viral markers in the patients’ blood were tested. The serum HBsAg was quantified using the Abbott RT-PCR HBV surface antigen test with a minimum detection limit of 0.05 IU/ml. The Hepatitis-B e antigen (HBeAg) was measured by Abbott Laboratories real-time PCR (Roche Diagnostics). All 15 subjects in this study were negative in HBeAg. The serum HBV DNA was tested by Roche Diagnostics COBAS amplifier / COBAS TaqMan HBV test (Roche Diagnostics), with the minimum detection limit of 20 IU/ml. Nine of the 15 individuals (60%) in this study had HBV DNA above the detection line before the therapy. In addition, blood samples from four CHB patients were collected before and 6 h after Pegylated interferon α-2b injection alone (180 μg, qw) in the Affiliated Infectious Diseases Hospital of Soochow University. The medical ethics committee of the Affiliated Infectious Diseases Hospital of Soochow University gave ethical approval for this work, and written informed consent was obtained from all subjects.

### Mice

C57BL/6 mice were purchased from Laboratory Animal Center of Soochow University and all *in vivo* experimental procedures in mice have been approved by the Committee for Animal Welfare at Soochow University. 6-8 weeks old mice were used in most of experiments. All protocols and procedures for mouse study were performed in accordance with the Laboratory Animal Management Regulations. IFNα/γ-R^-/-^ mice were nice gifts from Dr. Sudan He (Center of Systems Medicine Chinese Academy of Medical Sciences, Institute of Systems Medicine). *Ifnar1^-/-^* mice were nice gifts from Dr. Chunsheng Dong (Soochow University). *Stat1^-/-^* mice were nice gifts from Dr. Eugene Y. Chinn (Soochow University).

### Animal experiments

For mouse experiments with Aspirin treatment, Aspirin was first prepared with DMSO (final concentration: 5%) and then diluted in PBS containing 5% (vol/vol) Cremophor EL (Macklin, cat #C804845). Aspirin and the control were intraperitoneally (*i.p.*) injected into mice. After 2 h, the mice were injected with mouse IFNβ (1,000 IU/g, *i.p.*). For mouse experiments with Salicylate treatment, Salicylate was diluted in PBS and injected into mice (*i.p.*) at 200 μg/g. After 12 h, mice were infected with VSV (1×10^8^ PFU per gram body mouse, *i.p.*) for 24 h. The mice were euthanized and the tissues from all the groups were collected and ground.

### Cell lines and cell culture

HEK293T, A549, Raw264.7, HeLa, HT1080, THP1 and HepG2 cells were obtained from ATCC. 2fTGH cells were kindly provided by Dr. Serge Y. Fuchs (University of Pennsylvania). THP1 cells were cultured in RPMI 1640 medium (HyClone) and other cells above were cultured in Dulbecco’s modified Eagle’s medium (DMEM; HyClone) supplemented with 10% FBS (GIBCO, Life Technologies), 100 units/ml penicillin and 100 μg/ml streptomycin. All cells were cultured at 37°C under 5% CO_2_.

### Expression constructs and reagents

Myc-PTP1B was purchased from YouBio, China. Myc-IRF9, Flag-IFNAR1, HA-IFNAR2 and HA-Ub were nice gifts from Dr. Serge Y. Fuchs from University of Pennsylvania, America. The HBV-1.3 construct was a gift from Prof. Xiaonan Zhang from Shanghai Public Health Clinical Center, China. Flag-GFP-IRF9 were generated using PCR amplified from Myc-tagged human IRF9. All mutations were generated by QuickChange site-Directed Mutagenesis Kit (Stratagene, 210518). All transient transfections were carried out using LongTrans (Ucallm) according to manufacturer’s instructions. The reagents in this study are listed here: Salicylate (Sigma, Cat #S2679), Aspirin for cells and mice (Solarbio, Cat #A8830), Aspirin for humans (Bayer S.p.A., HJ20160685), recombinant human IFNα (PEPROTECH, Cat #300-02A), recombinant mouse IFNβ (R&D systems, Cat #12401-1), PPi Cocktail (Selleck, Cat #B15002), PTP inhibitor I (Selleck, Cat #S3696) and TRIzol (TAKARA, Cat #9109).

### Chromatin immunoprecipitation and qPCR assay (ChIP-qPCR)

The cells were cross-linked by adding formaldehyde to final concentration of 1% and incubated at 37°C for 10 min. Crosslinking reactions were quenched with 0.125 M glycine for 5 min at room temperature. Cells were washed with PBS and resuspended in ChIP lysis buffer (1% SDS; 10 mM EDTA; 50 mM Tris-HCl, pH = 8.0) containing protease inhibitors (1 mM PMSF). Samples were then centrifuged at 13,000 rpm for 10 min to shear DNA to lengths between 200 and 1,000 base pairs. Sonicated lysates were then diluted with 1 ml ChIP dilution buffer (1% tritonX-100; 2 mM EDTA; 150 mM NaCl; 20 mM Tris-HCl, pH = 8.1). The anti-IRF9 antibody was added and incubated overnight on a rotor at 4°C. Protein G-agarose beads (16-266; Millipore) were washed twice and then added into the samples. The mixture was incubated for 3 h on a rotor at 4°C. Then, the mixture was washed sequentially with TSE I (0.1% SDS; 1% Triton X-100; 2 mM EDTA; 20 mM Tris-HCl, pH 8.1; 150 mM NaCl), TSE II (0.1% SDS; 1% Triton X-100; 2 mM EDTA; 20 mM Tris-HCl, pH 8.1; 500 mM NaCl), Buffer III (0.25 M LiCl; 1% NP40; 1% deoxycholate; 10 mM Tris-HCl, pH 8.1; 1 mM EDTA) and TE (10 mM Tris-HCl, pH 8.0 and 1 mM EDTA, pH 8.0). The histone complex from the beads was eluted by adding 0.5 ml Extraction Solution (1% SDS and 0.1 M NaHCO_3_). Then 20 μl of 5 M NaCl were added to the combined eluates and reverse histone-DNA crosslinks were performed by heating at 65°C overnight. The samples were treated with RNase A and proteinase K, and then Immunoprecipitated DNA and the input DNA were purified. Immunoprecipitated DNA was analyzed with absolute RT-qPCR. The amplification product was expressed as a percentage of the input for each condition.

### TCID_50_ assay

Cells were infected with VSV for 24 h. VSV viral titers were determined by a standard 50% tissue culture-infective dose (TCID_50_) assay. First, the supernatants containing VSV were collected and the supernatants were serially diluted with DMEM. Then the supernatants were put on the monolayer of Vero cells in 96-well plates. The TCID_50_ was calculated by the Spearman-Karber algorithm.

### Screening of small molecule compound library

The drug library with 214 of FDA-approved small molecule compounds from plant sources was purchased from Selleck Chemicals (L2000-Z659310). All drugs were diluted to 1 mM using DMSO. Cells were plated in 12-well plates and treated with IFNα (1,000 IU/ml) together with DMSO (Ctrl) or each compound (1 μM) for 12 h. Cells were harvested and lysed. Then the protein levels of pY112-IRF9 and IRF9 were determined by Western Blot.

### CRISPR-Cas9-mediated genome editing

The lentiCRISPRv2 vector was a gift from Dr. Fangfang Zhou (Soochow University, China). Small guide RNAs that target the gene (IRF9-KO-F: 5’-CACCGCCTGGACAGCAACTCAGGA-3’, IRF9-KO-R: 5’-AAACTCCTGAGT TGCTGTCCAGGC-3’) were cloned into the lentiCRISPRv2 vector and were then transfected into cells. Forty-eight hours after transfection, the cells were cultured under puromycin selection (1.5 μg/ml) for two weeks. The knockout effect was identified by Western Blot or RT-qPCR analysis. After that, cells were transferred to 96-well plates and cultured for further experiments.

### Mass spectrometry analysis (MS)

HEK293T cells were transfected with F-G-IRF9, and harvested in lysis buffer containing 150 mM NaCl, 20 mM Tris-HCl (pH7.4), 1% Nonidet P-40, 0.5 mM EDTA, PMSF (50 μg/ml) and protease inhibitors mixtures (Sigma). The proteins were subjected to SDS-PAGE gels and minimally stained with Coomassie brilliant blue (Beyotime, P0017S). The gel bands with target proteins were excised, and then were digested with trypsin. The resulting tryptic peptides were purified with C18 Zip Tip and then analyzed by an Orbitrap Elite hybrid mass spectrometer (Thermo Fisher) coupled with a Dionex LC.

### Viral infection *in vitro*

Before infection, cells were washed and cultured in serum-free medium. Then cells were infected with Vesicular stomatitis virus (VSV), VSV-GFP, Sendai virus (SeV), Influenza A Virus (H1N1, PR/8/34), or Herpes simplex virus (HSV) for 1.5 h. After that, the supernatant was removed and cells were cultured in 10% FBS medium.

### Viral infection *in vivo*

For viral infection *in vivo*, 6-8 weeks old mice were injected intraperitoneally (*i.p.*) with VSV (1×10^8^ PFU per gram body mouse). After 24 h of infection, the lung, liver, spleen, kidney, heart tissues from each mouse were collected and ground into pieces for analysis by western blot or RT-qPCR.

### Flow cytometry (FCM)

The cell surface staining of IFNAR1 in PBMCs was performed using PE conjugated anti-IFNAR1-PE (Invitrogen, MA5-23630). Briefly, anti-IFNAR1-PE antibodies were diluted by 1 × PBS (1:100). Cell staining was performed at 4 °C for 30 min and acquired with BD FACS Canto II. FACS data were analyzed using FlowJo software (FlowJo).

### ISRE luciferase reporter gene assay

To detect IFNα-induced transcriptional activity, cells were transfected with the ISRE-Luciferase together with Renilla plasmids. After 36 h, cells were treated with or without Salicylate. The luciferase activity was measured using the Dual-Luciferase reporter assay system (E1910; Promega).

### Molecular Dynamics Simulation

The molecular dynamics (MD) simulations were performed using the software package GROMACS 2020.7. The simulation system employed the AMBER14SB force field, with a temperature of 300 K and a simulation time of 150 ns. After loading the protein, hydrogen atoms were added to complete the structure. The PTP1B-salicylic acid complex was dissolved in TIP3P water, with a minimum distance of 1.0 nm from the edges of the simulation box. Sodium ions were added to neutralize the system. The TIP3P explicit water model was selected, and periodic boundary conditions were set. The simulations followed a standard procedure including energy minimization, NVT equilibration, NPT equilibration, and production molecular dynamics simulations. The system was energy minimized using the steepest descent method for 50,000 steps. A 50,000-step NVT equilibration was performed with position restraints, at a temperature of 300 K with a timestep of 2 fs. A 50,000-step NPT equilibration was conducted with position restraints, at a temperature of 300 K with a timestep of 2 fs. A production molecular dynamics simulation of 150 ns was carried out in the NPT ensemble, with a timestep of 2 fs. Trajectory data was saved every 100 ps. Throughout the simulation, periodic boundary conditions were applied, and the LINCS algorithm was used to constrain all hydrogen-containing bonds. The temperature and pressure were maintained using the Parrinello-Rahman algorithm, with a temperature coupling constant of 0.1 ps and a pressure coupling constant of 1.0 ps. Long-range electrostatic interactions were calculated using the PME algorithm.

### Immunoprecipitation

Cells were harvested in lysis buffer containing 150 mM NaCl, 20 mM Tris-HCl (pH7.4), 1% Nonidet P-40, 0.5 mM EDTA, PMSF (50 μg/ml) and protease inhibitors mixtures (Sigma). Then the supernatant was collected after centrifugation. The specific antibodies were added into the supernatant and mixed overnight on a rotor at 4 °C. Protein G-agarose beads (16-266; Millipore) were washed twice and then added into the supernatant. The mixture was incubated for 3 h on a rotor at 4 °C. For immunoprecipitation of Flag-tagged proteins, the anti-Flag affinity gel (Selleck) or M2 affinity gel (A2220; Sigma-Aldrich) was added to the whole cell lysates and incubated for 3 h on a rotor at 4°C. Then the mixture was centrifuged at 4 °C and the supernatant was discarded. Then the precipitates were washed three times by washing buffer and were analyzed by Western Blot.

### Western Blot

Cells were harvested in lysis buffer containing 150 mM NaCl, 20 mM Tris-HCl (pH7.4), 1% Nonidet P-40, 0.5 mM EDTA, PMSF (50 μg/ml) and protease inhibitors mixtures (Sigma). Phosphatase inhibitor cocktail (PPi cocktail, Selleck, Cat #B15002, A, B tube, 100×) was added into the above lysis buffer when the phosphorylation of proteins was detected. PPi cocktail A, which was dissolved in water, contains 100 mM Sodium Fluoride, 100 mM Sodium Orthovanadate, 400 mM Sodium Tartrate, 115 mM Sodium Molybdate and 200 mM Imidazole. PPi cocktail B, which was dissolved in DMSO, contains 2.5 mM (-)-p-Bromotetramisole oxalate, 500 µM Cantharidin and 500 nM Microcystin LR (from Microcystis aeruginosa). Equivalent amounts of proteins were subjected to SDS-PAGE and transferred to polyvinylidene difluoride membranes (Millipore). The membranes were blocked with 5% nonfat milk or 5% BSA for 0.5 h at room temperature and then incubated with the primary antibodies (Abs) overnight at 4°C. After washing three times with PBST, the membranes were incubated with the secondary Abs (HRP-conjugated goat anti-rabbit or goat anti-mouse [Bioworld]) for 1 h at room temperature. After washing three times with PBST, the membranes were visualized with ECL Prime (Thermo Scientific). The densities of Western blot were analyzed by the ImageJ program. The antibody against phosphorylated IRF9 at Y112 was generated by GL Biochem (Shanghai, China) through immunization of rabbits with GRMDVAEPYKV (pY) QLL-Cys peptide. The validation of this antibody is presented in Figure 2E and S1E.

The antibodies in this study are listed here: Anti-IRF9 (Santa Cruz, Cat #sc-365893), Anti-pan-pY (Cell Signalling Technology, Cat #8954), Anti-Flag (Sigma, Cat #F7425), Anti-Tubulin (Proteintech, Cat #66031-1-Ig), Anti-GFP (Santa Cruz, Cat #sc-9996), Anti-p-STAT1(Y701) (Cell Signalling Technology, Cat #9167), Anti-STAT1 (Cell Signalling Technology, Cat #9176), Anti-PTP1B (Proteintech, Cat #11334-1-AP), Anti-β-Actin (Proteintech, Cat #66009), Anti-Myc (Abmart, Cat #M20002), Anti-HA (Abcam, Cat #ab9110), Anti-IFNAR1-PE (Invitrogen, Cat #MA5-23630), Anti-IFNAR1 (Sino Biological, Cat #13222-T20), Anti-β-Trcp (Cell Signalling Technology, Cat #11984),Anti-pan-AcK (Santa Cruz, Cat #sc-32268), Anti-Viperin (Abcam, Cat #107359), Anti-IFIT1 (Santa Cruz, Cat #sc-134948), Anti-HA (H1N1) (Sino Biological, Cat #11684-T56), Anti-p-STAT1 (S727) (Cell Signalling Technology, Cat #8826), Anti-p-STAT2 (Y690) (Cell Signalling Technology, Cat #88410), Anti-VSV-G (Abcam, Cat #ab1874), Anti-PKR (Cell Signalling Technology, Cat #3072S) and Anti-GAPDH (Goodhere Biological Technology, Cat #AB-M-M001).

### Real-time quantitative PCR (RT-qPCR)

Cells were harvested with TRIzol reagent (TAKARA). The cDNA was synthesized from 1 µg of total RNA using the All in one 5× RT MasterMix (abm, #G592) and subjected to RT-qPCR with different primers in the presence of SYBR Green Supermix (Selleck) using a StepOne Plus real-time PCR system (Applied Bioscience). Quantitation of all target gene expression was normalized to the control genes Actin (human) or GAPDH (mouse). All data from the cells in experimental groups were shown as fold changes normalized to that in either unstimulated or uninfected cells accordingly. The results were analyzed from at least three biological duplication and were shown as the average mean ± SD. The primer sequences are as following: human

human *Ifit1* (5’-CACAAGCCATTTTCTTTGCT-3’ and 5’-ACTTGGCTGCATATC GAAAG-3’);

CHIP-*Ifit1* (5’-CATTGGGTTTCTGCAGCACT-3’ and 5’-GTGTAAGCTGTGGGT GTGTC-3’);

CHIP-Isg54 (5’-ACCAGGCCGATGAAACATCCCT-3’ and 5’-ACAAGTGGCC TCTGGTTCCT-3’);

CHIP-*Viperin* (5’-AGAAACCAGGAATGCTCGCCCCG-3’ and 5’-TCTGAGCAA CCTGTCATTGGGGA -3’);

Actin (5’-ACCAACTGGGACGACATGGAGAAA-3’ and 5’-ATAGCACAGCCT GGATAGCAACG-3’)

GAPDH (5’-GGCCTTCCGTGTTCCTACC-3’ and 5’-AGCCCAAGATGCCCTTCAGT - 3’);

human *Isg54* (5’-GGAGAGCAATCTGCGACAG-3’ and 5’-GCTGCCTCAT TTAGACCTCTG -3’);

human *Isg15* (5’-GGGACCTGACGGTGAAGATG-3’ and 5’-CGCCGATCTT CTGGGTGAT-3’);

HBV DNA (5’-GAGTGTGGATTCGCACTCC-3’ and 5’-GAGGCGAGGGAGTT CTTCT -3’);

human *Ifnar1* (5’-CGCCTGTGATCCAGGATTATCC-3’ and 5’-TGGTGTGTG CTCTGGCTTTCAC-3’);

human *Parp11* (5’-CACACTGGGAGAATGTGAATACTC-3’ and 5’-CGACTG GAATAAGCAGCATCTC-3’);

human *β-Trcp* (5’-GGACACAAACGAGGCATTGCCT-3’ and 5’-CAACGCACCA ATTCCTCATGGC-3’);

mouse *Ifit1* (5’-GCCTATCGCCAAGATTTAGATGA-3’ and 5’-TTCTGGATTT AACCGGACAGCA-3’);

mouse *Isg15* (5’-GGTGTCCGTGACT AACTCCAT-3’ and 5’-CTGTACCACTA GCATCACTGTG -3’);

mouse *Mx1* (5’-GACCATAGGGGTCTTGACCAA-3’ and 5’-AGACTTGCTCTTT CTGAAAAGCC-3’);

human *Ifnβ* (5’-CATTACCTGAAGGCCAAGGA-3’ and 5’-CAGCATCTGCTGGT TGAAGA-3’);

SeV (5’-GATGACGATGCCGCAGCAGTAG-3’ and 5’-CCTCCGATGTCAGTTGGTTCAC TC-3’);

VSV (5’-ACGGCGTACTTCCAGATGG-3’ and 5’-CTCGGTTCAAGATCCAGG T-3’);

H1N1 (5’-TTCTAACCGAGGTCGAAACG-3’ and 5’-ACAAAGCGTCTACGCTGCAG-3’);

HSV-UL42 (5’-CCAACGCCAAGACGGTGTA-3’ and 5’-GGGGGTCGTGA GGAAGAAC-3’);

HSV-ICP27 (5’-ATCGCACCTTCTCTGTGGTC-3’ and 5’-GCAAATCTTCT GGGGTTTCA-3’);

### Peripheral Blood Mononuclear Cell (PBMC) extraction

After obtaining the patient’s consent, 10 ml of whole blood was collected before and 6 h after treatment. Fresh anticoagulated whole blood was collected and diluted with an equal volume of 1 × PBS. Then 5 ml of human lymphocyte separation medium (DAKEWE, cat #7111011) was added into the tube containing whole blood. The volume ratio of human lymphocyte separation medium, whole blood and 1 × PBS was 1: 1: 1. The samples were centrifuged at 700 g for 20 min with slow acceleration and deceleration. The middle layer of tunica albuginea (PBMCs) were taken at the end of centrifugation. The PBMCs were washed with 1 × PBS and then resuspended in cell saving medium (NCM, cat #C40050) and frozen in -80°C.

### Statistical analysis

Data are shown as means ± SD and comparison between different groups was analyzed using two-tailed unpaired Student’s *t*-test. Values of p < 0.05 were considered statistically significant. *p < 0.05, **p < 0.01, ***p < 0.001; NS, not significant. The results of RT-qPCR were performed in Graph Pad Prism.

## Data availability

All data generated or analysed during this study are included in Figs. 1–8 and Supplementary Figs. 1–8. The data that support the findings of this study are available from the corresponding author on reasonable request.

## REFERENCES

Bianconi, V., Violi, F., Fallarino, F., Pignatelli, P., Sahebkar, A., and Pirro, M. (2020). Is Acetylsalicylic Acid a Safe and Potentially Useful Choice for Adult Patients with COVID-19? Drugs 80, 1383–1396.

Buster, E.H., Flink, H.J., Cakaloglu, Y., Simon, K., Trojan, J., Tabak, F., So, T.M., Feinman, S.V., Mach, T., Akarca, U.S., et al. (2008). Sustained HBeAg and HBsAg loss after long-term follow-up of HBeAg-positive patients treated with peginterferon alpha-2b. Gastroenterology 135, 459–467.

Elwood, P., Morgan, G., Watkins, J., Protty, M., Mason, M., Adams, R., Dolwani, S., Pickering, J., Delon, C., and Longley, M. (2024). Aspirin and cancer treatment: systematic reviews and meta-analyses of evidence: for and against. British journal of cancer 130, 3–8.

Flink, H.J., van Zonneveld, M., Hansen, B.E., de Man, R.A., Schalm, S.W., and Janssen, H.L. (2006). Treatment with Peg-interferon alpha-2b for HBeAg-positive chronic hepatitis B: HBsAg loss is associated with HBV genotype. The American journal of gastroenterology 101, 297–303.

Forner, A., Reig, M., and Bruix, J. (2018). Hepatocellular carcinoma. Lancet (London, England) 391, 1301–1314.

Guo, T., Zuo, Y., Qian, L., Liu, J., Yuan, Y., Xu, K., Miao, Y., Feng, Q., Chen, X., Jin, L., et al. (2019). ADP-ribosyltransferase PARP11 modulates the interferon antiviral response by mono-ADP-ribosylating the ubiquitin E3 ligase β-TrCP. Nature microbiology 4, 1872–1884.

Iftikhar, U., Ahmed, M., Saeed, H., Saleem, N., Bashir, A., Farooq, M.S., Kamran, S.H., and Saleem, Z. (2020). Comparative safety and efficacy of conventional interferon versus pegylated-interferon based therapy for HCV: A retrospective cohort study from Gujranwala, Pakistan. Pakistan journal of pharmaceutical sciences 33, 2037–2045.

Ignat, M.D., Balta, A.A.S., Barbu, R.E., Draganescu, M.L., Nechita, L., Voinescu, D.C., Nechita, A., Stefanopol, I.A., Busila, C., and Baroiu, L. (2024). Antiviral Therapy of Chronic Hepatitis B Virus between Present and Future. Journal of clinical medicine 13.

Jeng, W.J., Papatheodoridis, G.V., and Lok, A.S.F. (2023). Hepatitis B. Lancet (London, England) 401, 1039–1052.

Kumar, K.G., Krolewski, J.J., and Fuchs, S.Y. (2004). Phosphorylation and specific ubiquitin acceptor sites are required for ubiquitination and degradation of the IFNAR1 subunit of type I interferon receptor. The Journal of biological chemistry 279, 46614–46620.

Kumar, K.G., Tang, W., Ravindranath, A.K., Clark, W.A., Croze, E., and Fuchs, S.Y. (2003). SCF(HOS) ubiquitin ligase mediates the ligand-induced down-regulation of the interferon-alpha receptor. The EMBO journal 22, 5480–5490.

Lam, R., and Lim, J.K. (2024). Advances in discovery of novel investigational agents for functional cure of chronic hepatitis B: A comprehensive review of phases II and III therapeutic agents. World journal of hepatology 16, 331–343.

Liao, C.L., Lin, Y.L., Wu, B.C., Tsao, C.H., Wang, M.C., Liu, C.I., Huang, Y.L., Chen, J.H., Wang, J.P., and Chen, L.K. (2001). Salicylates inhibit flavivirus replication independently of blocking nuclear factor kappa B activation. Journal of virology 75, 7828–7839.

Lucifora, J., Xia, Y., Reisinger, F., Zhang, K., Stadler, D., Cheng, X., Sprinzl, M.F., Koppensteiner, H., Makowska, Z., Volz, T., et al. (2014). Specific and nonhepatotoxic degradation of nuclear hepatitis B virus cccDNA. Science (New York, NY) 343, 1221–1228.

Marcellin, P., Bonino, F., Yurdaydin, C., Hadziyannis, S., Moucari, R., Kapprell, H.P., Rothe, V., Popescu, M., and Brunetto, M.R. (2013). Hepatitis B surface antigen levels: association with 5-year response to peginterferon alfa-2a in hepatitis B e-antigen-negative patients. Hepatology international 7, 88–97.

Mazur, I., Wurzer, W.J., Ehrhardt, C., Pleschka, S., Puthavathana, P., Silberzahn, T., Wolff, T., Planz, O., and Ludwig, S. (2007). Acetylsalicylic acid (ASA) blocks influenza virus propagation via its NF-kappaB-inhibiting activity. Cellular microbiology 9, 1683–1694.

Millman, I., Loeb, L.A., Bayer, M.E., and Blumberg, B.S. (1970). Australia antigen (a hepatitis-associated antigen): purification and physical properties. The Journal of experimental medicine 131, 1190–1199.

Nan, J., Wang, Y., Yang, J., and Stark, G.R. (2018). IRF9 and unphosphorylated STAT2 cooperate with NF-κB to drive IL6 expression. Proceedings of the National Academy of Sciences of the United States of America 115, 3906–3911.

Qiu, Y., Su, M., Liu, L., Tang, Y., Pan, Y., and Sun, J. (2021). Clinical Application of Cytokines in Cancer Immunotherapy. Drug design, development and therapy 15, 2269–2287.

Rehermann, B., Lau, D., Hoofnagle, J.H., and Chisari, F.V. (1996). Cytotoxic T lymphocyte responsiveness after resolution of chronic hepatitis B virus infection. The Journal of clinical investigation 97, 1655–1665.

Roth, G.J., Stanford, N., and Majerus, P.W. (1975). Acetylation of prostaglandin synthase by Aspirin. Proceedings of the National Academy of Sciences of the United States of America 72, 3073–3076.

Song, A., Lin, X., Lu, J., Ren, S., Cao, Z., Zheng, S., Hu, Z., Li, H., Shen, C., and Chen, X. (2021). Pegylated Interferon Treatment for the Effective Clearance of Hepatitis B Surface Antigen in Inactive HBsAg Carriers: A Meta-Analysis. Frontiers in immunology 12, 779347.

Stark, G.R., and Darnell, J.E., Jr. (2012). The JAK-STAT pathway at twenty. Immunity 36, 503–514.

Tang, Y., Liang, H., Zeng, G., Shen, S., and Sun, J. (2022). Advances in new antivirals for chronic hepatitis B. Chinese medical journal 135, 571–583.

Trott, O., and Olson, A.J. (2010). AutoDock Vina: improving the speed and accuracy of docking with a new scoring function, efficient optimization, and multithreading. Journal of computational chemistry 31, 455–461.

van Bömmel, F., Stein, K., Heyne, R., Petersen, J., Buggisch, P., Berg, C., Zeuzem, S., Stallmach, A., Sprinzl, M., Schott, E., et al. (2023). A multicenter randomized-controlled trial of nucleos(t)ide analogue cessation in HBeAg-negative chronic hepatitis B. Journal of hepatology 78, 926–936.

Wiede, F., Lu, K.H., Du, X., Zeissig, M.N., Xu, R., Goh, P.K., Xirouchaki, C.E., Hogarth, S.J., Greatorex, S., Sek, K., et al. (2022). PTP1B Is an Intracellular Checkpoint that Limits T-cell and CAR T-cell Antitumor Immunity. Cancer discovery 12, 752–773.

Zheng, H., Gupta, V., Patterson-Fortin, J., Bhattacharya, S., Katlinski, K., Wu, J., Varghese, B., Carbone, C.J., Aressy, B., Fuchs, S.Y., et al. (2013). A BRISC-SHMT complex deubiquitinates IFNAR1 and regulates interferon responses. Cell reports 5, 180–193.

Zheng, H., Qian, J., Baker, D.P., and Fuchs, S.Y. (2011a). Tyrosine phosphorylation of protein kinase D2 mediates ligand-inducible elimination of the Type 1 interferon receptor. The Journal of biological chemistry 286, 35733–35741.

Zheng, H., Qian, J., Varghese, B., Baker, D.P., and Fuchs, S. (2011b). Ligand-stimulated downregulation of the alpha interferon receptor: role of protein kinase D2. Molecular and cellular biology 31, 710–720.

Zuo, Y., Feng, Q., Jin, L., Huang, F., Miao, Y., Liu, J., Xu, Y., Chen, X., Zhang, H., Guo, T., et al. (2020). Regulation of the linear ubiquitination of STAT1 controls antiviral interferon signaling. Nature communications 11, 1146.

Zuo, Y., He, J., Liu, S., Xu, Y., Liu, J., Qiao, C., Zang, L., Sun, W., Yuan, Y., Zhang, H., et al. (2022). LATS1 is a central signal transmitter for achieving full type-I interferon activity. Science advances 8, eabj3887.

