## Supplementary material for "Aspirin improves both reactivity and durability of type-I interferon signaling to achieve functional cure of chronic hepatitis B": Suppl Figs and Legends

S1.

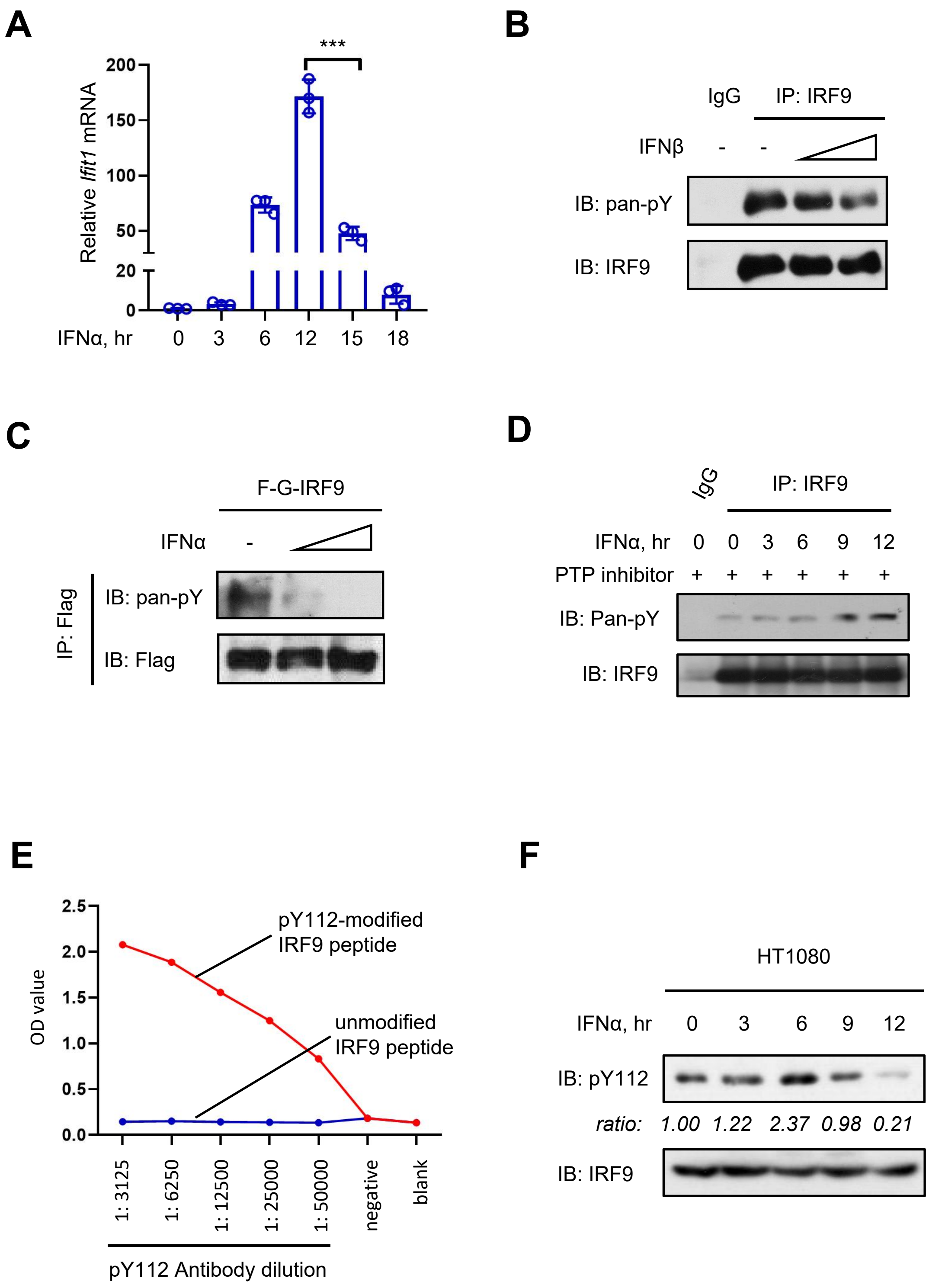
A
***
Relative Ifit1 mRNA
200
150
100
50
20
0
IFNα, hr
0
3
6
12
15
18
B
IgG
IP: IRF9
IFNβ
-
-
IB: pan-pY
IB: IRF9
C
F-G-IRF9
IFNα
-
IP: Flag
IB: pan-pY
IB: Flag
D
IgG
IP: IRF9
IFNα, hr
0 0 3 6 9 12
PTP inhibitor
+ + + + + +
IB: Pan-pY
IB: IRF9
E
OD value
2.5
2.0
1.5
1.0
0.5
0.0
pY112-modified
IRF9 peptide
unmodified
IRF9 peptide
1: 3125
1: 6250
1: 12500
1: 25000
1: 50000
negative
blank
pY112 Antibody dilution
F
HT1080
IFNα, hr
0 3 6 9 12
IB: pY112
ratio:
1.00 1.22 2.37 0.98 0.21
IB: IRF9

**Figure S1. IRF9 tyrosine phosphorylation is reduced during IFN-I treatment**

(A) RT-qPCR analysis of *Ifit1* mRNA in HEK293T cells treated with IFNα (1,000 IU/ml) for indicated times.

(B) Immunoprecipitation (IP)-immunoblotting (IB) analysis of pan-tyrosine phosphorylation (pan-pY) of IRF9 in Raw264.7 cells treated with IFNβ (300 and 600 IU/ml) for 12 h.

(C) IP-IB analysis of pan-pY of IRF9 in HT1080 cells transfected with Flag-GFP-tagged IRF9 (F-G-IRF9) and treated with IFNα (1,000 and 3,000 IU/ml) for 12 h.

(D) IP-IB analysis of pan-pY of IRF9 in HEK293T cells treated with PTP inhibitor I (10 μM) and IFNα (1,000 IU/ml) for indicated times.

(E) ELISA analysis of the binding of the pY112-IRF9 antibody to either pY112-modified IRF9 peptides or unmodified IRF9 peptides.

(F) Western blot analysis of pY112-IRF9 levels in HT1080 cells treated with IFNα (1,000 IU/ml) for indicated times.

***$p < 0.001$ (two-tailed unpaired Student's *t*-test). Data are shown as means ± SD of three biological replicates (A), or are representative of three independent experiments (B-D and F).

S2.

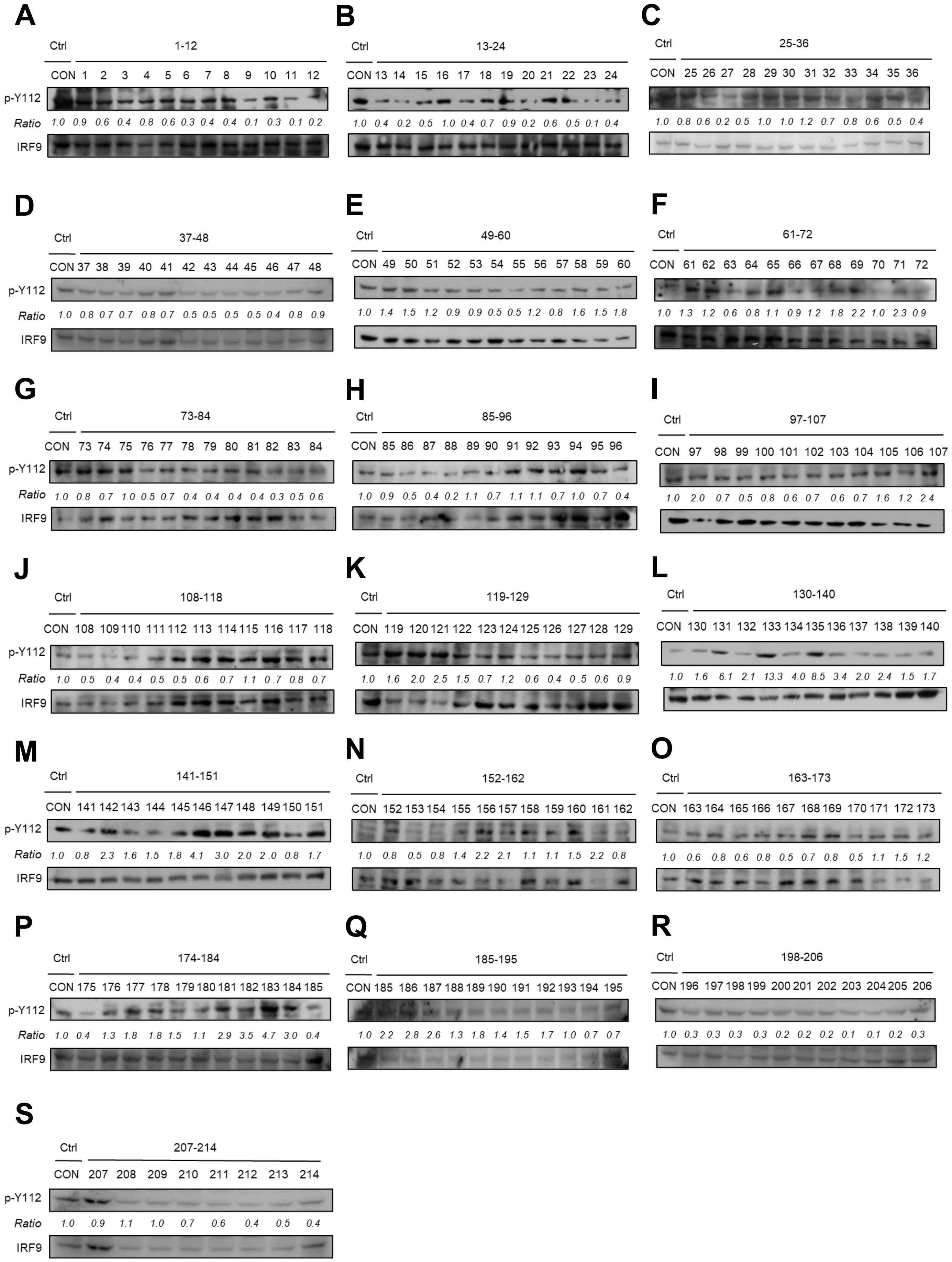
A
Ctrl 1-12
CON 1 2 3 4 5 6 7 8 9 10 11 12
p-Y112
Ratio 1.0 0.9 0.6 0.4 0.8 0.6 0.3 0.4 0.4 0.1 0.3 0.1 0.2
IRF9
B
Ctrl 13-24
CON 13 14 15 16 17 18 19 20 21 22 23 24
1.0 0.4 0.2 0.5 1.0 0.4 0.7 0.9 0.2 0.6 0.5 0.1 0.4
C
Ctrl 25-36
CON 25 26 27 28 29 30 31 32 33 34 35 36
1.0 0.8 0.6 0.2 0.5 1.0 1.0 1.2 0.7 0.8 0.6 0.5 0.4
D
Ctrl 37-48
CON 37 38 39 40 41 42 43 44 45 46 47 48
p-Y112
Ratio 1.0 0.8 0.7 0.7 0.8 0.7 0.5 0.5 0.5 0.5 0.4 0.8 0.9
IRF9
E
Ctrl 49-60
CON 49 50 51 52 53 54 55 56 57 58 59 60
1.0 1.4 1.5 1.2 0.9 0.9 0.5 0.5 1.2 0.8 1.6 1.5 1.8
F
Ctrl 61-72
CON 61 62 63 64 65 66 67 68 69 70 71 72
1.0 1.3 1.2 0.6 0.8 1.1 0.9 1.2 1.8 2.2 1.0 2.3 0.9
G
Ctrl 73-84
CON 73 74 75 76 77 78 79 80 81 82 83 84
p-Y112
Ratio 1.0 0.8 0.7 1.0 0.5 0.7 0.4 0.4 0.4 0.4 0.3 0.5 0.6
IRF9
H
Ctrl 85-96
CON 85 86 87 88 89 90 91 92 93 94 95 96
1.0 0.9 0.5 0.4 0.2 1.1 0.7 1.1 1.1 0.7 1.0 0.7 0.4
I
Ctrl 97-107
CON 97 98 99 100 101 102 103 104 105 106 107
1.0 2.0 0.7 0.5 0.8 0.6 0.7 0.6 0.7 1.6 1.2 2.4
J
Ctrl 108-118
CON 108 109 110 111 112 113 114 115 116 117 118
p-Y112
Ratio 1.0 0.5 0.4 0.4 0.5 0.5 0.6 0.7 1.1 0.7 0.8 0.7
IRF9
K
Ctrl 119-129
CON 119 120 121 122 123 124 125 126 127 128 129
1.0 1.6 2.0 2.5 1.5 0.7 1.2 0.6 0.4 0.5 0.6 0.9
L
Ctrl 130-140
CON 130 131 132 133 134 135 136 137 138 139 140
1.0 1.6 6.1 2.1 13.3 4.0 8.5 3.4 2.0 2.4 1.5 1.7
M
Ctrl 141-151
CON 141 142 143 144 145 146 147 148 149 150 151
p-Y112
Ratio 1.0 0.8 2.3 1.6 1.5 1.8 4.1 3.0 2.0 2.0 0.8 1.7
IRF9
N
Ctrl 152-162
CON 152 153 154 155 156 157 158 159 160 161 162
1.0 0.8 0.5 0.8 1.4 2.2 2.1 1.1 1.1 1.5 2.2 0.8
O
Ctrl 163-173
CON 163 164 165 166 167 168 169 170 171 172 173
1.0 0.6 0.8 0.6 0.8 0.5 0.7 0.8 0.5 1.1 1.5 1.2
P
Ctrl 174-184
CON 175 176 177 178 179 180 181 182 183 184 185
p-Y112
Ratio 1.0 0.4 1.3 1.8 1.8 1.5 1.1 2.9 3.5 4.7 3.0 0.4
IRF9
Q
Ctrl 185-195
CON 185 186 187 188 189 190 191 192 193 194 195
1.0 2.2 2.8 2.6 1.3 1.8 1.4 1.5 1.7 1.0 0.7 0.7
R
Ctrl 198-206
CON 196 197 198 199 200 201 202 203 204 205 206
1.0 0.3 0.3 0.3 0.3 0.2 0.2 0.2 0.1 0.1 0.2 0.3
S
Ctrl 207-214
CON 207 208 209 210 211 212 213 214
p-Y112
Ratio 1.0 0.9 1.1 1.0 0.7 0.6 0.4 0.5 0.4
IRF9

**Figure S2. Identification of the small molecules that upregulate pY112-IRF9 in IFN-I signaling**

(A-S) A drug library containing 214 clinically approved small molecules from plant sources was employed. Western blot was performed to analyze pY112-IRF9 levels using a specific anti-pY112-IRF9 antibody in HEK293T cells treated with 214 small molecular compounds (1 mM) each, together with IFNα (1,000 IU/ml) for 12 h.

Data are representative of three independent experiments (A-S).

S3.

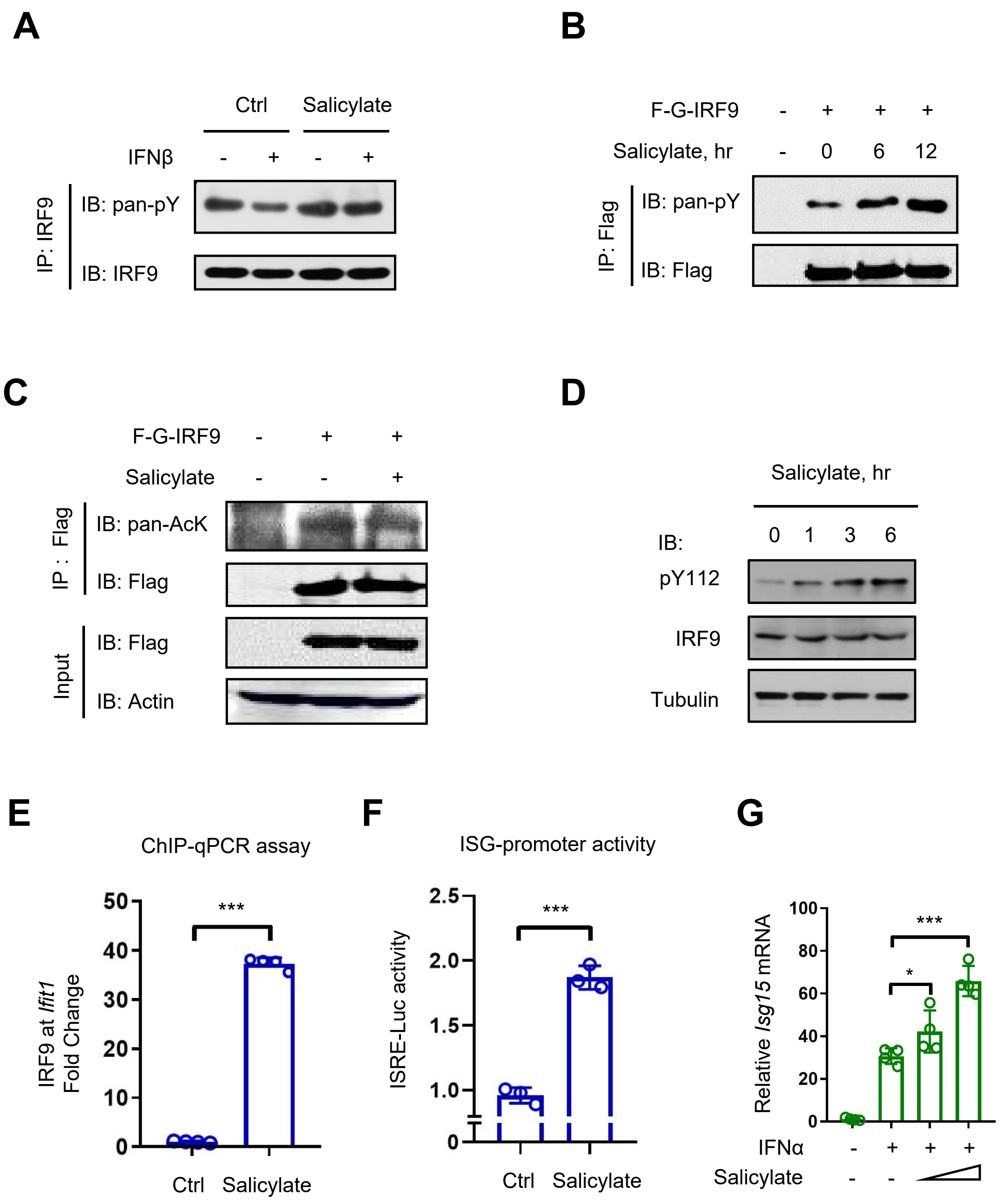

A
Ctrl
Salicylate
IFNβ - + - +
IP: IRF9
IB: pan-pY
IB: IRF9
B
F-G-IRF9 - + + +
Salicylate, hr - 0 6 12
IP: Flag
IB: pan-pY
IB: Flag
C
F-G-IRF9 - + +
Salicylate - - +
IP : Flag
IB: pan-AcK
IB: Flag
Input
IB: Flag
IB: Actin
D
Salicylate, hr
0 1 3 6
IB:
pY112
IRF9
Tubulin
E
ChIP-qPCR assay
IRF9 at Ifit1
Fold Change
0
10
20
30
40
50
***
Ctrl
Salicylate
F
ISG-promoter activity
ISRE-Luc activity
0
1.0
1.5
2.0
2.5
***
Ctrl
Salicylate
G
Relative Isg15 mRNA
0
20
40
60
80
100
***
*
IFNα - + + +
Salicylate - -

**Figure S3. Salicylate upregulates pY112-IRF9 levels and promotes IRF9-ISRE binding**

(A) IP-IB analysis of pan-pY of IRF9 in Raw264.7 cells pre-treated with PBS (Ctrl) or Salicylate (100 μg/ml, 6 h) and then treated with IFNβ (300 IU/ml, 12 h).

(B) IP-IB analysis of pan-pY of IRF9 in HEK293T cells transfected with F-G-IRF9 and then treated with Salicylate (100 μg/ml) for 6 and 12 h.

(C) IP-IB analysis of pan-lysine acetylation (AcK) of IRF9 in HEK293T cells transfected with F-G-IRF9 and then treated with or without Salicylate (100 μg/ml, 6 h).

(D) Western blot analysis of pY112-IRF9 in HEK293T cells treated with Salicylate (100 μg/ml) for indicated times.

(E) ChIP-qPCR analysis of the binding of cellular IRF9 proteins to the ISRE of the representative ISG (*Ifit1*) gene in 2fTGH cells treated with Salicylate (100 μg/ml, 6 h).

(F) ISRE-Luciferase activity in HEK293T cells transfected with the ISRE-Luciferase and Renilla reporters and then treated with or without Salicylate (100 μg/ml) for 24 h.

(G) RT-qPCR analysis of *Isg15* mRNA in THP1 cells pre-treated with Salicylate (300 and 600 μg/ml, 2 h) and then treated with IFNα (1,000 IU/ml, 6 h).

*$p < 0.05$, ***$p < 0.001$ (two-tailed unpaired Student's *t*-test). Data are shown as means ± SD of at least three biological replicates (E-G) , or are representative of three independent experiments (A-D).

S4.

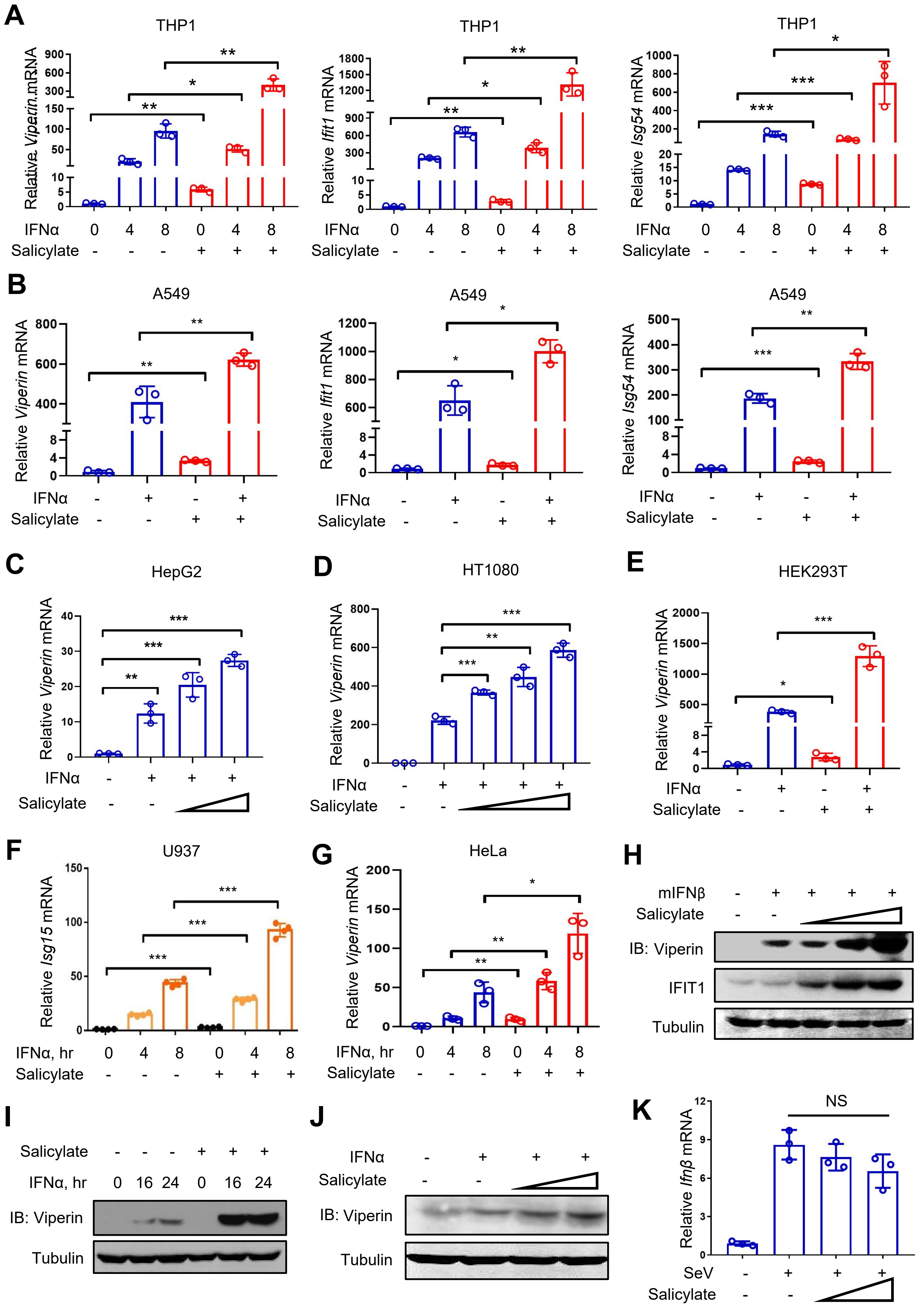
A
THP1
Relative Viperin mRNA
Relative Ifit1 mRNA
Relative Isg54 mRNA
IFNα 0 4 8 0 4 8
Salicylate - - - + + +
B
A549
Relative Viperin mRNA
Relative Ifit1 mRNA
Relative Isg54 mRNA
IFNα - + - +
Salicylate - - + +
C
HepG2
Relative Viperin mRNA
IFNα - + + +
Salicylate - -
D
HT1080
Relative Viperin mRNA
IFNα - + + + +
Salicylate - -
E
HEK293T
Relative Viperin mRNA
IFNα - + - +
Salicylate - - + +
F
U937
Relative Isg15 mRNA
IFNα, hr 0 4 8 0 4 8
Salicylate - - - + + +
G
HeLa
Relative Viperin mRNA
IFNα, hr 0 4 8 0 4 8
Salicylate - - - + + +
H
mIFNβ - + + + +
Salicylate - -
IB: Viperin
IFIT1
Tubulin
I
Salicylate - - - + + +
IFNα, hr 0 16 24 0 16 24
IB: Viperin
Tubulin
J
IFNα - + + +
Salicylate - -
IB: Viperin
Tubulin
K
NS
Relative Ifnβ mRNA
SeV - + + +
Salicylate - -

**Figure S4. Salicylate enhances IFN-I Signaling**

(A and B) RT-qPCR analysis of *Viperin*, *Ifit1, Isg54* mRNA in THP1 (A) and A549 (B) cells pre-treated with Salicylate (100 μg/ml, 6 h) and then treated with IFNα (1,000 IU/ml) as indicated (A) or for 6 h (B).

(C and D) RT-qPCR analysis of *Viperin* mRNA in HepG2 (C) and HT1080 (D) cells pre-treated with Salicylate (100 and 300 μg/ml, 6 h) and then treated with IFNα (1,000 IU/ml) for 6 h.

(E-G) RT-qPCR analysis of the representative ISG mRNA in HEK293T (E), U937 (F) and HeLa (G) cells pre-treated with Salicylate (100 μg/ml, 6 h) and then treated with IFNα (1,000 IU/ml) as indicated.

(H) Western blot analysis of Viperin and IFIT1 proteins in Raw264.7 cells pre-treated with increased amounts of Salicylate for 6 h and then treated with IFNβ (300 IU/ml, 6 h).

(I and J) Western blot analysis of Viperin in THP1 (I) and 2fTGH (J) cells pre-treated with increased amounts of Salicylate (100, 200 μg/ml, 6 h) and then treated with IFNα (1,000 IU/ml) as indicated.

(K) RT-qPCR analysis of *Ifnβ* mRNA in HT1080 cells pre-treated with Salicylate (100 and 300 μg/ml, 6 h) and then infected with SeV (MOI = 1.0) for 8 h.

NS, not significant ($p > 0.05$), *$p < 0.05$, **$p < 0.01$ , ***$p < 0.001$ (two-tailed unpaired Student's *t*-test). Data are shown as means ± SD of at least three biological replicates (A-G, K), are representative of three independent experiments (H-J).

S5.

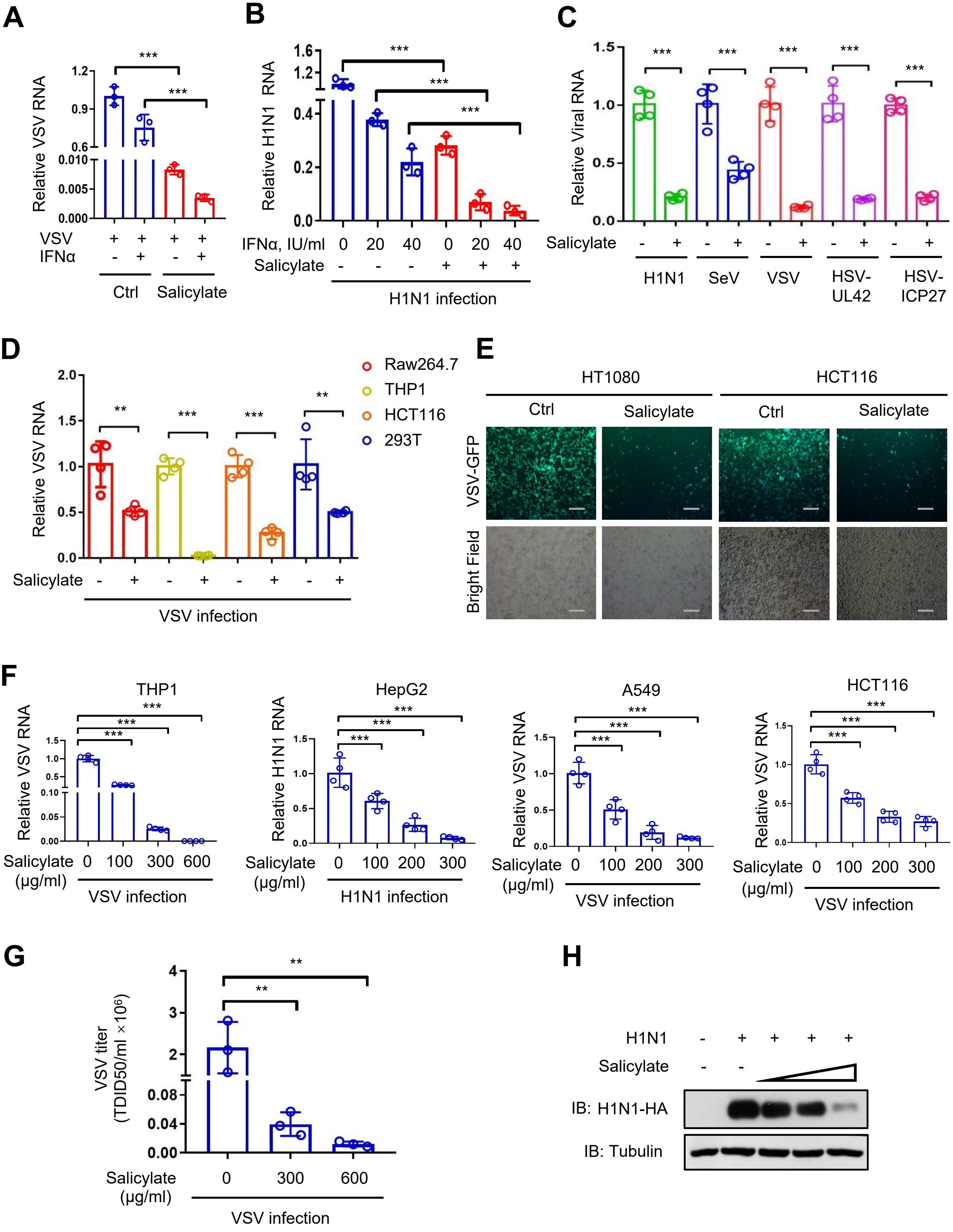
A
Relative VSV RNA
VSV + + + +
IFNα - + - +
Ctrl
Salicylate
B
Relative H1N1 RNA
IFNα, IU/ml 0 20 40 0 20 40
Salicylate - - - + + +
H1N1 infection
C
Relative Viral RNA
Salicylate - + - + - + - + - +
H1N1
SeV
VSV
HSV-UL42
HSV-ICP27
D
Relative VSV RNA
Raw264.7
THP1
HCT116
293T
Salicylate - + - + - + - +
VSV infection
E
HT1080
HCT116
Ctrl
Salicylate
VSV-GFP
Bright Field
F
THP1
HepG2
A549
HCT116
Relative VSV RNA
Relative H1N1 RNA
Salicylate (μg/ml) 0 100 300 600
Salicylate (μg/ml) 0 100 200 300
VSV infection
H1N1 infection
G
VSV titer (TDID50/ml ×10⁶)
Salicylate (μg/ml) 0 300 600
VSV infection
H
H1N1 - + + + +
Salicylate - -
IB: H1N1-HA
IB: Tubulin

**Figure S5. Salicylate promotes IFN-I antiviral immune activity**

(A) RT-qPCR analysis of VSV RNA in 2fTGH cells treated with IFNα (20 IU/ml) for 24 h, together with Salicylate (100 μg/ml), and then infected with VSV (MOI = 1.0) for 24 h.

(B) RT-qPCR analysis of H1N1 RNA in Raw264.7 cells treated with IFNα and Salicylate as (A), and then infected with H1N1 (MOI = 1.0) for 24 h.

(C) RT-qPCR analysis of Viral RNA in A549 cells treated with Salicylate (300 μg/ml) and then immediately infected with H1N1, SeV, VSV or HSV (MOI = 1.0) for 24 h.

(D) RT-qPCR analysis of VSV in Raw264.7, THP1, HCT116 and HEK293T cells treated with Salicylate and VSV as (C).

(E) VSV-GFP viruses were observed by fluorescence in HT1080 and HCT116 cells treated with Salicylate and VSV-GFP as (C). Scale bars, 100 μm.

(F and G) RT-qPCR analysis of Viral RNA in THP1, HepG2, A549 and HCT116 cells (F) or VSV titers in HEK293T cells (G) treated with Salicylate as indicated and then immediately infected with viruses as (C).

(H) Western blot analysis of H1N1-encoded HA proteins in HEK293T cells treated with Salicylate and H1N1 as (C).

NS, not significant ($p > 0.05$), *$p < 0.05$, **$p < 0.01$ , ***$p < 0.001$ (two-tailed unpaired Student's *t*-test). Data are shown as means ± SD of at least three biological replicates (A-D, F, G), are representative of three independent experiments (H).

S6.

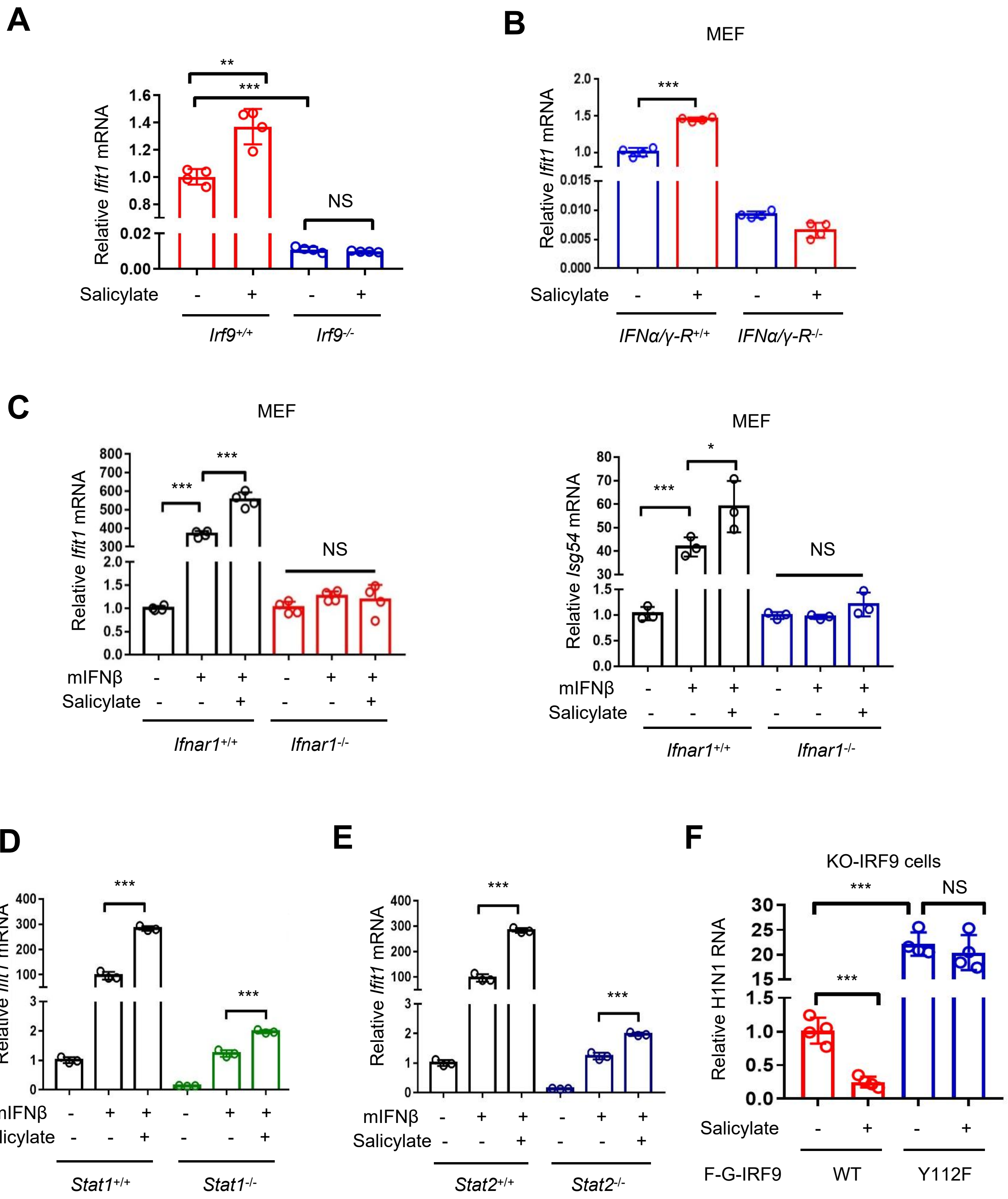

A
Relative Ifit1 mRNA
Salicylate
Irf9+/+
Irf9-/-
NS
B
MEF
Relative Ifit1 mRNA
Salicylate
IFNα/γ-R+/+
IFNα/γ-R-/-
C
MEF
Relative Ifit1 mRNA
mIFNβ
Salicylate
Ifnar1+/+
Ifnar1-/-
NS
MEF
Relative Isg54 mRNA
mIFNβ
Salicylate
Ifnar1+/+
Ifnar1-/-
NS
D
Relative Ifit1 mRNA
mIFNβ
Salicylate
Stat1+/+
Stat1-/-
E
Relative Ifit1 mRNA
mIFNβ
Salicylate
Stat2+/+
Stat2-/-
F
KO-IRF9 cells
Relative H1N1 RNA
NS
Salicylate
F-G-IRF9
WT
Y112F

**Figure S6. Salicylate-mediated enhancement of IFN-I signaling is dependent of IRF9**

(A) RT-qPCR analysis of *Ifit1* mRNA in *Irf9*$^{+/+}$ and *Irf9*$^{-/-}$ HEK293T cells treated with or without Salicylate (300 μg/ml) for 6 h .

(B) RT-qPCR analysis of *Ifit1* mRNA in *IFNα/γ-R*$^{+/+}$ and *IFNα/γ-R*$^{-/-}$ MEF cells treated with or without Salicylate as (A).

(C) RT-qPCR analysis of *Ifit1* and *Isg54* mRNA in *Ifnar1*$^{+/+}$ and *Ifnar1*$^{-/-}$ MEF cells pre-treated with Salicylate (100 μg/ml, 6 h) and then treated with IFNβ (1,000 IU/ml) for 6 h.

(D and E) RT-qPCR analysis of *Ifit1* mRNA in *Stat1*$^{+/+}$ and *Stat1*$^{-/-}$ (D) or *Stat2*$^{+/+}$ and *Stat2*$^{-/-}$ (E) cells treated with Salicylate and IFNβ as (C).

(F) IRF9-WT or IRF9-Y112F was stably expressed in IRF9 knockout (KO) cells. RT-qPCR was used to analyze H1N1 RNA levels in these cells treated with Salicylate (300 μg/ml) and then immediately infected with H1N1 (MOI = 1.0) for 24 h. NS, not significant ($p > 0.05$), *$p < 0.05$, **$p < 0.01$, ***$p < 0.001$ (two-tailed unpaired Student's *t*-test). Data are shown as means ± SD of at least three biological replicates (A-F).

S7.

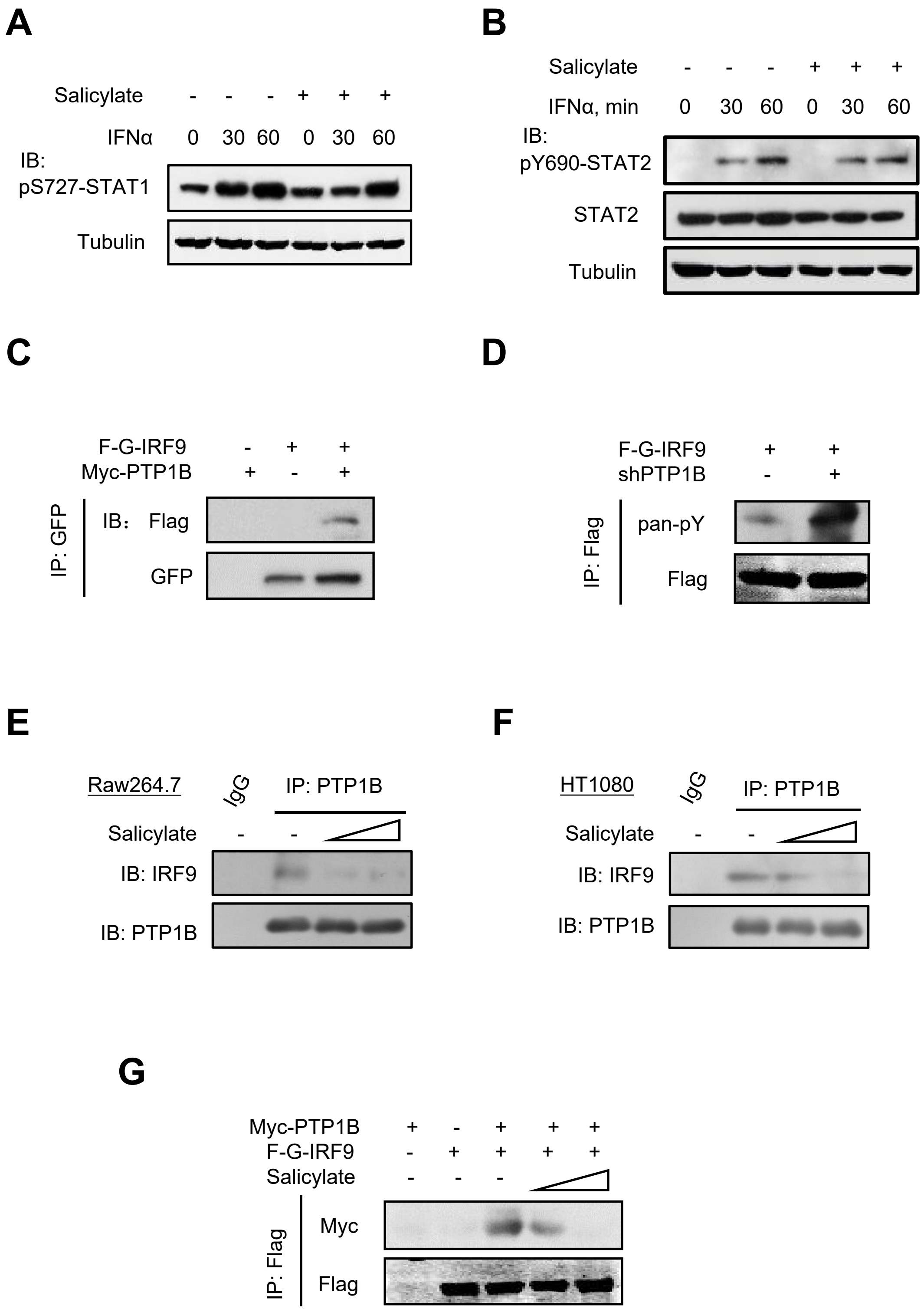

A
Salicylate - - - + + +
IFNα 0 30 60 0 30 60
IB: pS727-STAT1
Tubulin
B
Salicylate - - - + + +
IFNα, min 0 30 60 0 30 60
IB: pY690-STAT2
STAT2
Tubulin
C
F-G-IRF9 - + +
Myc-PTP1B + - +
IP: GFP
IB: Flag
GFP
D
F-G-IRF9 + +
shPTP1B - +
IP: Flag
pan-pY
Flag
E
Raw264.7
IgG
IP: PTP1B
Salicylate - -
IB: IRF9
IB: PTP1B
F
HT1080
IgG
IP: PTP1B
Salicylate - -
IB: IRF9
IB: PTP1B
G
Myc-PTP1B + - + + +
F-G-IRF9 - + + + +
Salicylate - - -
IP: Flag
Myc
Flag

**Figure S7. PTP1B regulates IRF9 tyrosine phosphorylation, while Salicylate inhibits PTP1B-IRF9 interaction**

(A) Western blot analysis of pS727-STAT1 in A549 cells pre-treated with Salicylate (100 μg/ml, 6 h) and then treated with IFNα (1,000 IU/ml) for 30 and 60 min.

(B) Western blot analysis of pY690-STAT2 in HT1080 cells pre-treated with Salicylate (100 μg/ml, 6 h) and then treated with IFNα (1,000 IU/ml) for 30 and 60 min.

(C) IP-IB analysis of the interaction between F-G-IRF9 and Myc-PTP1B in HEK293T cells transfected with F-G-IRF9 and Myc-PTP1B.

(D) IP-IB analysis of pan-pY of IRF9 in HEK293T cells transfected with F-G-IRF9, together with control shRNAs (-) or shRNAs against PTP1B (shPTP1B).

(E and F) IP-IB analysis of the IRF9-PTP1B interaction in Raw264.7 (E) and HT1080 (F) cells treated with increased amounts of Salicylate (100 and 300 μg/ml) for 6 h.

(G) IP-IB analysis of the interaction between F-G-IRF9 and Myc-PTP1B in HEK293T cells treated with increased amounts of Salicylate (100 and 300 μg/ml) for 6 h.

Data are representative of three independent experiments (A-G).

S8.

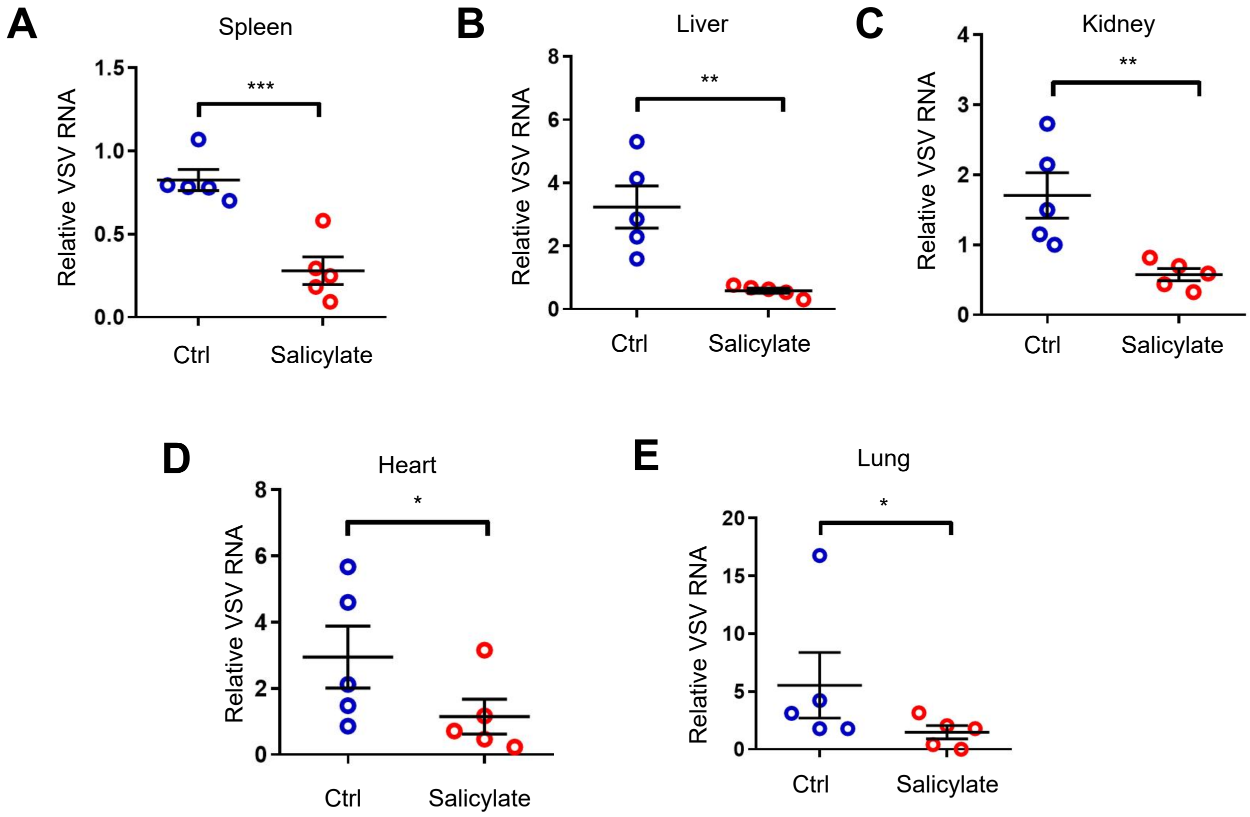
A
Spleen
Relative VSV RNA
1.5
1.0
0.5
0.0
***
Ctrl
Salicylate
B
Liver
Relative VSV RNA
8
6
4
2
0
**
Ctrl
Salicylate
C
Kidney
Relative VSV RNA
4
3
2
1
0
**
Ctrl
Salicylate
D
Heart
Relative VSV RNA
8
6
4
2
0
*
Ctrl
Salicylate
E
Lung
Relative VSV RNA
20
15
10
5
0
*
Ctrl
Salicylate

**Figure S8. Salicylate administration inhibits virus infection *in vivo***

(A-E) C57BL/6 mice ($n = 5$) were injected intraperitoneally (*i.p.*) with Salicylate (200 μg per gram body mouse). After 12 h, mice were infected with VSV ($1 \times 10^8$ PFU per gram body mouse, *i.p*) for 24 h. RT-qPCR was used to analyze VSV RNA levels in mouse spleen, liver, kidney, heart and lung tissues. $^*p < 0.05$, $^{**}p < 0.01$, $^{***}p < 0.001$ (two-tailed unpaired Student's *t*-test). All graphs show the means ± SEM for five individual mice (A-E).
